# The Registry of Pregnant Women at Cruces University Hospital: an ethical framework for prospective research with preanalytical optimization of maternal plasma processing

**DOI:** 10.64898/2026.07.17.26357942

**Authors:** Itziar Gonzalez-Moro, Haizea Sanchez-Garcia, Tatiana Medina Cuesta, Ane Rodríguez Liro, Maravillas del Pilar Espín López, Susana Esquivel González, Eider Quintana Ochoa de Alda, Mikel de la Peña-Sanz, Laura Marín Cano, Naia Sarasua-Blanco, Paula Ortiz Salinas, Abril Sanfeliu Padullés, Águeda Ruiz Adrián, Ainara Martínez Isidoro, Ainhoa Aldaiturriaga Otaola, Ainhoa Aramburu Gil, Alba García Gil, Alexandra Sáenz Sáenz, Almudena Heredia Campos, Amaia Fernández Salado, Ana Isabel Ramírez Jarana, Ana Isabel Tobar López, Ana José Casarojos Oses, Andrea Martínez de Marañón Toral, Andrea Satiago Hidalgo, Ane Silva Díaz, Anne Basterrechea Miguel, Aranzazu Castaños Lasa, Aúrea Esteras Vadillo, Beatriz González Díaz, Beatriz Lucio Moral, Begoña de Diego Ajenjo, Carolina Martín López, Celia González Rodríguez, Clara Navas Pariente, Claudia Guadalupe Toledo, Cristina Barriocanal Contreras, Elisabeth Castañeira Huerta, Esther Arana Arbide, Esther Mieza Vázquez, Estibaliz González de Audicana Aparicio, Garazi Meléndez Cárcamo, Idoia Zorrilla Padilla, Irati Herranz Ortega, Irati Pérez Pérez, Irati Saenz Martín, Iratxe Cruz Quijano, Isabel Rodríguez Pérez, Itziar Rebolledo Cantero, Janire Garcés Conde, Jasone de Dios Idígoras, Jesús Sánchez Martínez, Lara Martín Andrés, Leticia Álvarez Paule, Leire Valle Ruiz de Larrea, Lucía Ortiz Sosa, Lucía Salceda Couce, Lucía Vázquez Gómez, Maite Arechabaleta Basterrechea, Maite Terreros Salsidúa, María Cano Martín, María Fidalgo Astorquia, María Vázquez Bautista, Mariana Alcíbar Laraudogoitia, Marina Carbajo Martín, Marta Martínez Iturriaga, Mireia Bonilla García, Mónica Izquierdo Rodriguez, Mónica Santiago Vélez, Nahikari López Quintana, Naia Marcoida Pichel, Nekane del Olmo Cobos, Nerea Albillos Rodríguez, Nerea Elguezabal Acebedo, Nerea Muñoz Toyos, Noivé Abascal Gómez, Nuria Torres López, Olga Peña Ruiz, Patricia Sánchez Zamarro, Paula Gállego Ortega, Paula Ortega Serrano, Raquel López Rodríguez, Rocío Sutil Arenas, Sandra García Santamaría, Sandra Martín Martín, Sara Mohamedi Munarriz, Sara Moreno Fernández, Sara Ruz Moreno, Sara Urbaneja Martín, Sonia Álvarez Barreiro, Susana Tamayo Echevarría, Tania Mateo Urdiales, Verónica Gómez Sobradillo, Virginia Velaz Azkoiti, Ylenia Bidea Iturbe, Yolanda Mateos Ibáñez, Zaloa Zenarruzabeitia Ireguenpagate, Ziortza Corbacho Rodríguez, Jaione Olaizola Canseco, Amaia Aiartzabuena Agirrebeitia, Iraia Gacia-Santisteban, Carmen Osuna Sierra, Marta Martínez Gutiérrez, Raquel Coya Guerrero, Leire Rodríguez Gómez, Jose Ramon Bilbao, María Jesús Mulas Martín, Jorge Burgos, Nora Fernandez-Jimenez

## Abstract

**Background:** Prospective pregnancy registries and biobanking infrastructures are essential for future translational studies investigating maternal, placental and offspring health. However, circulating nucleic acid analyses are highly sensitive to preanalytical variability, particularly regarding blood-collection tube type and sample processing conditions. We established a prospective pregnancy registry and biobanking workflow at Cruces University Hospital and evaluated the impact of preanalytical variables on circulating cell-free DNA (cfDNA) and cell-free RNA (cfRNA) preservation in maternal plasma collected at delivery.

**Methods:** The Registry of Pregnant Women at Cruces University Hospital was designed as a prospective infrastructure integrating placental sampling, maternal blood collection and ethically controlled future access to maternal and offspring clinical data. Within this framework, peripheral blood samples from 50 women at delivery were simultaneously collected into EDTA, Norgen and Roche tubes. Plasma samples processed within or after 24 hours following collection underwent cfDNA/cfRNA extraction, electrophoretic profiling, fluorometric quantification and RT-qPCR analyses targeting different stress-related genes.

**Results:** By the end of June 2026, 1,127 women had been prospectively recruited into the registry, with 661 plasma samples, 637 serum samples and 858 sets of four placental biopsies collected, processed and stored in the Basque Biobank. In the preanalytical substudy, EDTA tubes yielded higher cfDNA concentrations, likely reflecting reduced cellular preservation and genomic DNA contamination. In contrast, Roche tubes showed superior cfRNA preservation, with higher cfRNA concentrations and more consistent detection of the characteristic 5S rRNA peak compared with EDTA and Norgen tubes. Processing delays beyond 24 hours reduced cfRNA concentration, while associations between circulating transcripts and gestational age were more consistently detectable in preservative-containing tubes.

**Conclusions:** Prospective infrastructures like ours offer strong foundation for large scale, long-term studies in the framework of the Developmental Origins of Health and Disease hypothesis. Technically, Roche tubes provided superior cfRNA preservation and enhanced sensitivity for detecting subtle biological associations, supporting the importance of standardized preanalytical workflows within prospective pregnancy biobanking resources.

## Introduction

Pregnancy represents a critical developmental window during which maternal, placental and fetal physiological processes may influence health trajectories across the lifespan. Within the framework of the Developmental Origins of Health and Disease (DOHaD) hypothesis, increasing evidence suggests that molecular alterations occurring during gestation can contribute not only to pregnancy complications, but also to long-term metabolic, cardiovascular, neurodevelopmental and respiratory outcomes in the offspring^1^. In this context, the placenta has emerged as a particularly informative organ for understanding prenatal susceptibility and developmental programming, as placental molecular patterns may capture early biological adaptations linked to later health trajectories. Recent work has further highlighted the potential of placental molecular profiling to identify biologically meaningful signatures associated with neurodevelopmental outcomes and early-life risk trajectories^2^.

In addition, the identification of other minimally invasive biomarkers, capable of capturing dynamic biological processes during pregnancy, has become a major objective in translational obstetric research. Circulating cell-free DNA (cfDNA) and RNA (cfRNA) obtained from maternal plasma have emerged as promising tools for the study of pregnancy physiology and placental function. Since circulating nucleic acids originate from maternal, placental and fetal tissues, they may reflect ongoing biological processes such as placental remodeling, oxidative stress, inflammatory activation and cellular turnover^3,4^. Recent studies have demonstrated the potential utility of cfDNA and cfRNA profiling for the identification of molecular signatures associated with adverse pregnancy outcomes, including preterm birth (PTB) and preeclampsia^5–8^. However, beyond their possible diagnostic value during gestation, circulating nucleic acids may also provide insight into subtle biological variability across gestation, and into risk trajectories associated with maternal and placental adaptation that could serve as biomarkers of future health outcomes.

Despite this potential, one of the major challenges for the implementation of circulating nucleic acid studies in pregnancy is the strong influence of preanalytical variability. Factors such as blood-collection tube chemistry, sample transport, processing delay and storage conditions may substantially alter cfDNA and cfRNA integrity, concentration and biological interpretability^9–11^. This issue is especially relevant for cfRNA analyses, due to the fragmented and highly labile nature of circulating RNA molecules. Inadequate stabilization may increase technical noise, promote contamination from cellular nucleic acids released *ex vivo* and ultimately reduce the capacity to detect biologically meaningful associations.

Prospective pregnancy registries and biobanking infrastructures constitute an essential platform for future translational and longitudinal studies integrating molecular, clinical and developmental data. However, these initiatives must operate under strict ethical and data-protection frameworks, particularly when future access to maternal or offspring clinical records requires additional study-specific ethical approvals and explicit participant consent. Under these conditions, the adoption of standardized and reproducible workflows for biological sample collection and processing becomes a critical first step for future clinically integrated research.

In this study, we describe the implementation of a prospective pregnancy registry at the Cruces University Hospital in the Basque Country, Spain. This registry, named *Registro de Mujeres Embarazadas del Hospital Universitario Cruces* (Registry of Pregnant Women at Cruces University Hospital), enables the systematic collection of placental biopsies and blood samples at delivery, as well as controlled access to maternal and offspring clinical records following the provision of informed consent by mothers and additional legal guardians when applicable. In this context, we carried out a preanalytical substudy evaluating the impact of blood-collection tube type and processing delay on cfDNA and cfRNA preservation. Specifically, we compared EDTA - containing tubes and commercially available nucleic acid stabilization tubes (from Norgen and Roche), tubes in terms of circulating nucleic acid recovery, electrophoretic quality and transcript detection. In addition, we explored whether improved cfRNA preservation enhances the ability to detect subtle biological associations in maternal plasma, using gestational age (GA) at delivery and selected circulating transcripts as proof-of-principle examples.

## Methods

### Cohort infrastructure and ethical governance

The Registry of Pregnant Women at Cruces University Hospital is a prospective pregnancy registry and biobanking infrastructure established at the Cruces University Hospital, a tertiary-care academic hospital integrated within the Basque Public Health Service (Osakidetza) in the Basque Country, Spain, that serves as a reference center for multiple highly specialized clinical areas, including pediatrics, reproductive medicine and several complex medical and surgical specialties^12^. The institution comprises 847 hospital beds, 33 operating rooms and more than 6,000 healthcare professionals, attending over 190,000 emergency visits and approximately 3,800 deliveries annually. In addition to its clinical activity, the hospital maintains a strong academic and translational research profile through its integration with the University of the Basque Country teaching programs and the Biobizkaia Health Research Institute.

The registry was designed as a long-term translational platform aimed at enabling future studies on maternal, placental and offspring health within the framework of the DOHaD hypothesis. The infrastructure allows the systematic prospective collection of biological material obtained at delivery, including maternal peripheral blood samples and placental biopsies, together with the possibility of future controlled linkage to maternal and offspring clinical records following study-specific ethical approval procedures.

Participant recruitment was carried out through the Obstetrics and Gynecology service at delivery. Pregnant women admitted to Cruces University Hospital for delivery were informed about the objectives and scope of the registry prior to inclusion. Written informed consent was obtained from participating mothers and, when applicable under legal and ethical requirements, from the additional legal guardian authorizing future controlled access to offspring clinical information. Consent procedures specifically addressed the collection, storage and future research use of biological samples, as well as the governance conditions regulating potential future access to clinical data.

The registry was specifically structured around a governance model prioritizing data protection, participant autonomy and ethically controlled clinical access. In accordance with applicable European and Spanish data-protection regulations, clinical information was not directly integrated into the research database at the time of recruitment. Instead, any future extraction or linkage to maternal or offspring clinical variables requires independent approval by the corresponding Research Ethics Committee and must remain consistent with the scope of the informed consent provided by participating families. This design was intended to ensure strict compliance with ethical and legal standards regarding sensitive maternal and pediatric health data.

All biological samples were pseudonymized at collection using unique registry identifiers. Sample processing, transport and storage workflows were standardized to minimize preanalytical variability and ensure traceability throughout the biobanking process. Maternal blood samples were collected at delivery and processed according to predefined protocols for plasma isolation and downstream circulating nucleic acid analyses. Placental biopsies were systematically obtained immediately after delivery following standardized sampling procedures designed to preserve tissue quality for future molecular analyses.

The registry was conceived as an expandable infrastructure supporting future longitudinal and multi-omics studies integrating circulating biomarkers, placental molecular profiling and developmental outcomes. The implementation of harmonized preanalytical workflows constituted a central objective of the platform from its inception, given the known sensitivity of circulating nucleic acid analyses to sample handling and processing conditions.

### Sample collection workflow

The Registry of Pregnant Women at Cruces University Hospital includes a standardized protocol for the collection of placental tissue biopsies and maternal blood samples at delivery. Midwives from the Births Unit are responsible for the implementation of the collection workflow, and nearly 100 midwives have already participated in the registry procedures.

For placental sampling, the placenta was positioned with the umbilical cord facing upwards. A sterile punch biopsy tool was placed approximately 5 cm from the cord insertion site, selecting an area with minimal vascularization. The punch was used to perforate the placental tissue, and the obtained biopsy was transferred into the collection tube using the punch ejector. This procedure was repeated until four tissue cores were collected in the same tube. The tube was then inverted gently to ensure appropriate mixing with the RNAlater preservative solution.

Placental biopsies were maintained upright at room temperature overnight to facilitate complete penetration of the preservative solution. Before freezing, the preservative was removed and tissue cores were washed in phosphate-buffered saline (PBS) within the same tube. The four tissue cores were subsequently transferred into a new tube labeled with the study-specific donor code. Samples were frozen horizontally at −80 °C without isopentane and, once completely frozen, stored vertically at −80 °C.

For maternal blood collection within the general registry workflow, one 10 mL EDTA tube for plasma isolation and one 5 mL serum tube were collected from each participant, ideally during the initial examination upon admission and before administration of medication whenever possible. All tubes were labeled with the patient identification code and placed in designated transport racks.

Placental biopsies, blood tubes and signed informed consent forms were stored in a dedicated locker located at the Births Unit. Hospital orderlies transported samples to the Basque Biobank twice daily (around 8:30 and 13:00) during working days. Upon arrival at the Basque Biobank, plasma was immediately isolated and placental biopsies were processed and frozen following the standardized workflow described above. This procedure ensured sample processing within the first 24 hours for most specimens, except for those collected during weekends or holidays. All samples were registered within the biobank management system, where a unique coded identifier was generated to ensure pseudonymization, traceability and controlled future access to clinical records. The link between donor codes and any personal identifiable information is kept strictly confidential and secure under biobank governance, ensuring that the identity of the donors remains fully protected from any unauthorized access or external parties.

### Preanalytical substudy

Within the framework of the registry, a preanalytical substudy was conducted to evaluate the impact of blood-collection tube type and processing delay on cfDNA and cfRNA preservation in maternal plasma. Peripheral blood samples were collected from 50 women delivering at the hospital across a range of GAs, yielding a total of 150 plasma samples (**Additional File 1**). Among these participants, 9 delivered before 266 gestational days (<38 weeks), whereas the remaining 41 delivered at term (>266 days; maximum 293 days).

Peripheral blood (10 mL) was simultaneously collected at delivery into three different commercial blood-collection tubes: EDTA tubes (BD Vacutainer® K2EDTA Tubes, BD, 366643), Norgen cf-DNA/cf-RNA Preservative Tubes (Norgen Biotek, Catalog number 63950) and Roche Cell-Free DNA Collection Tubes (Roche, Catalog number 07785666001) (**Figure 1**). For these particular women, no serum tube was collected.

**Figure 1.**
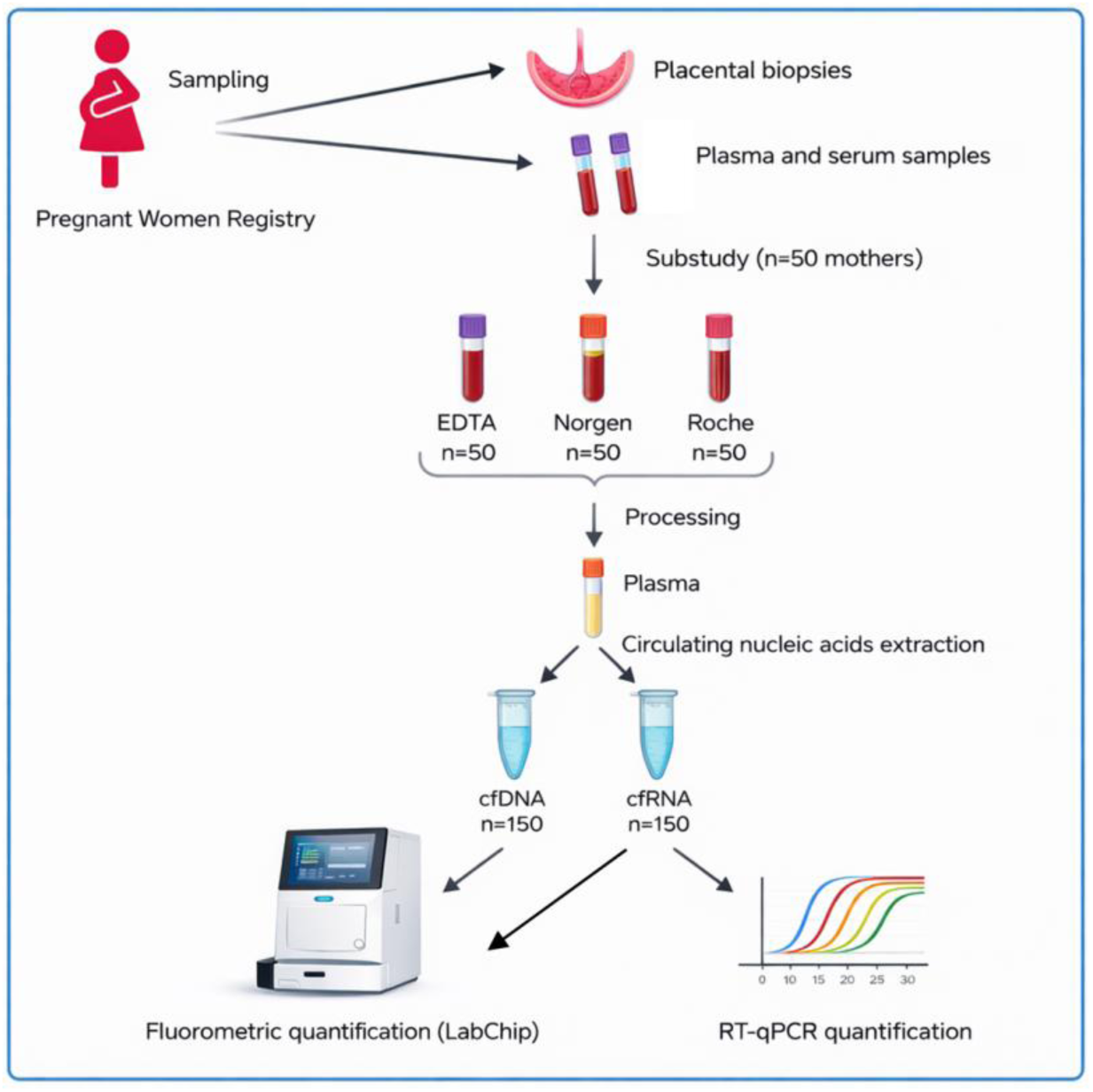
Study flowchart. Blood was drawn from 50 mothers participating in the Registry of Pregnant Women into different blood collection tubes (EDTA, Norgen, Roche). Samples were processed according to manufacturers’ recommendations and plasma was extracted. Circulating nucleic acids purification was performed to obtain cfDNA and cfRNA following the manufacturer’s protocol. The concentration of both cfDNA and cfRNA was calculated by fluorometric quantification and later by RT-qPCR.

Immediately after blood collection, tubes were gently inverted approximately 5–10 times to ensure adequate mixing of blood with the preservative solution. Samples were subsequently transported to the Basque Biobank following the routine registry workflow. Samples collected during weekdays and Sunday evenings were generally processed within the next 24 hours, whereas those collected from Friday afternoon onwards or during holidays were processed after more than 24 hours.

To ensure standardized handling across all participants, 50 preassembled collection kits were prepared in advance. Each kit contained the three blood-collection tubes together with written instructions reminding midwives to invert all tubes 5–10 times immediately after collection. No predefined collection order among tube types was established. Kits were used sequentially according to admission order until all 50 kits had been utilized.

Plasma isolation was performed according to the manufacturers’ recommendations. EDTA and Roche tubes were centrifuged at 1600 g for 10 min at room temperature, whereas Norgen tubes were centrifuged at 425 g for 20 min at room temperature. EDTA samples underwent an additional centrifugation step at 13000 g for 10 min at room temperature. For each tube, one 2 mL plasma aliquot was stored at −80 °C for the present substudy, while two additional 1 mL aliquots were stored for future analyses.

GA at delivery was retrieved from clinical records following approval by the corresponding Ethics Committee (**Additional File 1**).

### cfDNA/cfRNA analyses

Circulating cfDNA and cfRNA were simultaneously extracted from plasma using the Plasma/Serum cfc-DNA/cfc-RNA Advanced Fractionation Kit (Norgen Biotek, Catalog number 68300), following the manufacturer’s instructions. A spin column-based workflow including sequential binding, washing and elution steps was performed at room temperature. Plasma samples underwent lysis and fractionation through multiple centrifugation steps of variable speed to separately recover cfDNA and cfRNA fractions, while minimizing genomic DNA contamination. cfDNA and cfRNA were eluted separately into 20 μL final volumes and stored at −80 °C until analysis.

Both cfDNA and cfRNA were quantified fluorometrically using the LabChip GX Touch 24 Nucleic Acid Analyzer (Revvity, CLS138162). For cfDNA quantification, 1 μL of sample was used, whereas cfRNA quantification was performed using 2 μL of sample.

Electrophoretic profiles were evaluated to assess fragment size distribution and nucleic acid quality. cfDNA profiles were characterized by the expected mono-nucleosomal peak around ∼160–180 bp together with additional higher-order fragments, whereas cfRNA profiles were evaluated according to the presence and quality of the characteristic 5S rRNA peak and overall fragment distribution patterns.

### RT-qPCR

cfRNA samples were thawed on ice and analyzed by one-step RT-qPCR reactions in a final reaction volume of 10 μL using the ZymoScriptTM One-Step RT-qPCR Kit (Zymo Research, R3014), following the manufacturer’s instructions.

Expression levels of *GDF15* and *ARHGEF28* were determined using *RPLP0* (ribosomal protein lateral stalk subunit P0) as the endogenous control. Each sample was analyzed in duplicate, and each reaction contained 2 μL of cfRNA. Primer sequences are listed in **Table 1**.

**Table 1.**
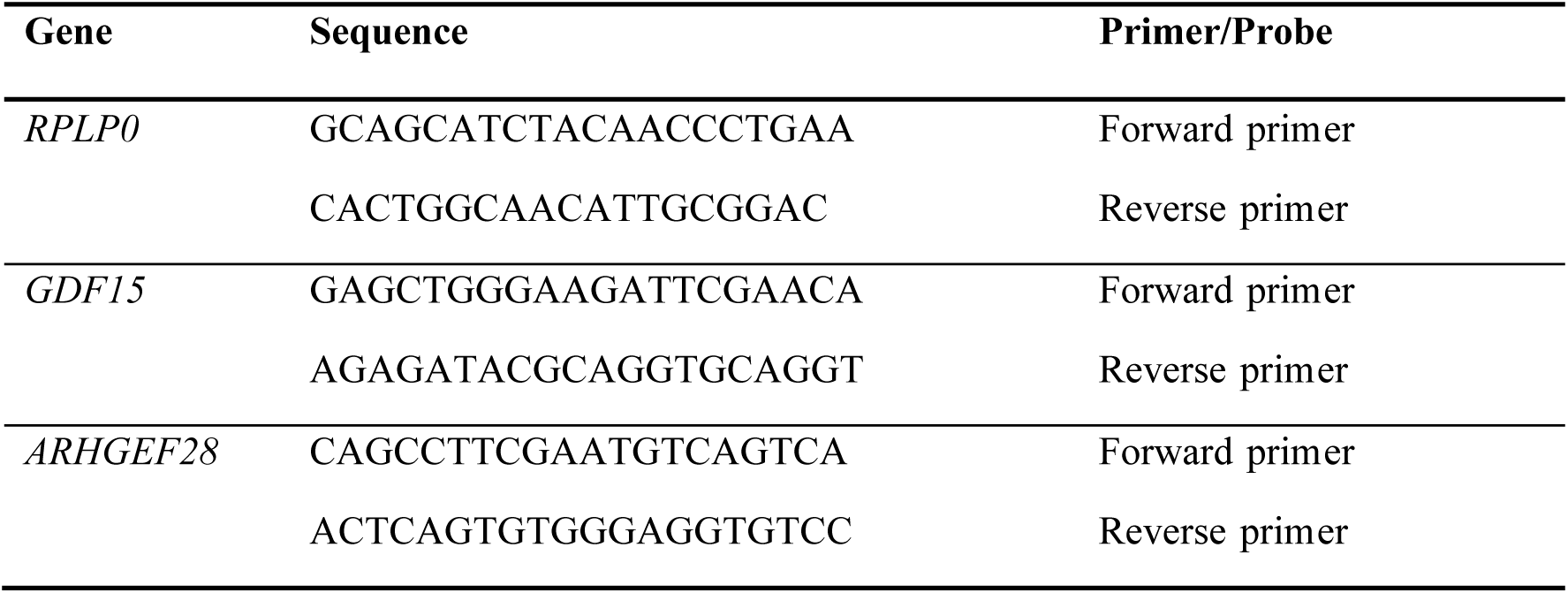
Primer sequences used for RT-qPCR.

RT-qPCR was performed using the following thermal cycling conditions: 50 °C for 10 min for cDNA synthesis, 95 °C for 10 min for initial denaturation, followed by 42 amplification cycles consisting of 95 °C for 10 s and 60 °C for 30 s. For SYBR Green -based assays, melting-curve analysis was subsequently performed from 65 °C to 95 °C with 0.5 °C increments every 5 s to confirm appropriate PCR product formation.

Quantification cycle (Cq) values were obtained using Bio-Rad CFX Maestro 1.0 software (version 4.0.2325.0418). Mean Cq values from duplicate reactions were used for downstream analyses. Samples considered undetected were excluded from further analyses. The expression of *GDF15* and *ARHGEF28* was normalized using the average Cq value of the endogenous control *RPLP0* and relative expression was calculated using the 2^−ΔΔCq^ method^13^.

### Statistics

Pearson correlation coefficients were used to evaluate linear relationships between quantitative variables. Student’s t-tests were used for pairwise comparisons, and repeated-measures analysis of variance (ANOVA) was applied to assess differences across blood-collection tube types. Statistical significance was defined as p < 0.05 (*), p < 0.01 (**) and p < 0.001 (***). Data are presented as mean ± standard deviation (SD).

## Results

### Registry recruitment and sample collection

By the end of June 2026, 1,127 women had been prospectively enrolled in the Registry of Pregnant Women at Cruces University Hospital. Among them, 1,099 had singleton pregnancies, while 28 had multiple pregnancies. The first mother was enrolled on the study June 2025. As part of the standardized collection workflow, 661 plasma samples, 637 serum samples and 858 sets of 4 placental biopsies were collected. All plasma samples were processed within 24 hours from blood collection in EDTA tubes. Samples exceeding 24 hours from collection were not processed further (except in the particular cases of the substudy). Mean maternal age at delivery was 34.36 ± 5.02 years, and mean GA at delivery was 39.31 ± 1.42 weeks. Among the newborns, 54.2% were female and 45.8% were male.

These results demonstrate the feasibility of integrating standardized maternal blood and placental tissue collection into routine obstetric clinical practice while maintaining controlled ethical and preanalytical conditions. At the time of publication of this manuscript, the recruitment is still ongoing, with no predefined end date.

### Blood-collection tube type and processing delay influence cfDNA preservation

Analysis of cfDNA extracted from maternal blood revealed differences in cfDNA concentration across the three blood collection tube types. Electrophoretic profiles from the LabChip system showed the characteristic cfDNA fragmentation pattern, with a dominant mono-nucleosomal peak centered around ∼160-180 bp and additional higher-order peaks corresponding to di- and tri-nucleosomal fragments (∼300-600 bp) (**Figure 2A**, **Additional File 2**). A broader distribution of longer fragments extending up to ∼3000 bp was also detected, and shorter fragments within the 100-150 bp range were also observed, consistent with the expected variability in cfDNA fragmentation.

**Figure 2.**
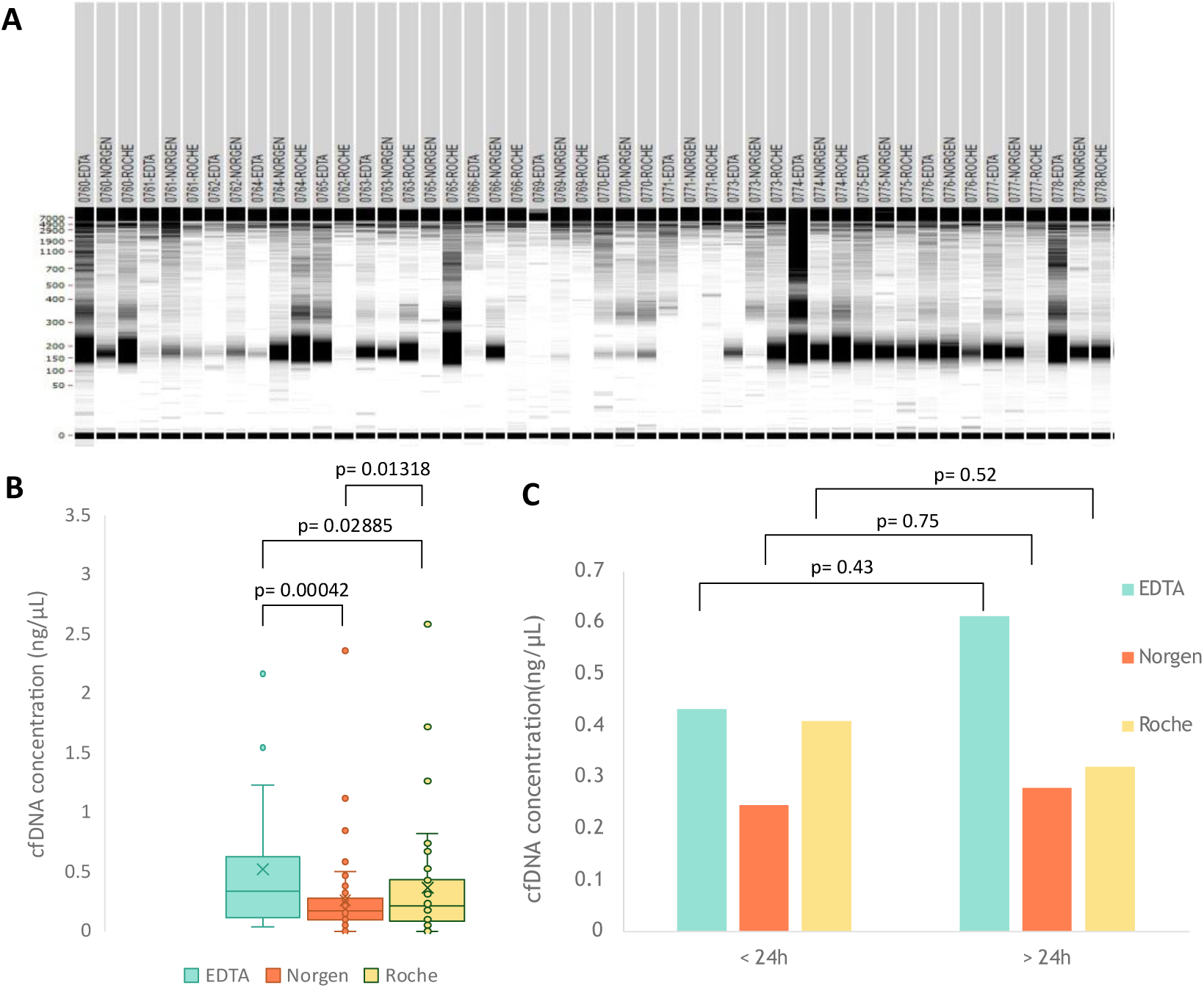
Analysis of cfDNA yield and fragment distribution across blood-collection tubes. **A.** Representative electrophoretic profiles obtained by Labchip analysis showing cfDNA fragment size distribution for samples across the three blood-collection tubes (EDTA, Norgen and Roche). **B.** cfDNA concentration (ng/µL) quantified within the 100-3000 bp range. A repeated-measures ANOVA (subjects = 50 women; factor = tube type) revealed a significant effect of tube type on cfDNA yield (F( 2, 98) = 9.142, p = 0.00023). Post-hoc paired comparisons showed that EDTA tubes yielded significantly higher cfDNA concentrations than Norgen (p = 0.00042). Significant differences were also observed between EDTA and Roche (p = 0.02885), as well as between Norgen and Roche (p = 0.01318). **C.** cfDNA concentration(ng/µL) stratified by processing time (< 24 h *vs.* > 24 h) and grouped by tube type. Post-hoc tests showed no significant differences between early and delayed processing for any tube type (EDTA: p = 0.43; Norgen: p= 0.75; Roche p = 0.52), indicating that processing delay had minimal impact on cfDNA yield under the conditions tested.

cfDNA quantification analysis within the 100–3000 bp range demonstrated a significant effect of blood-collection tube type on cfDNA yield (**Figure 2B**). Repeated-measures ANOVA (subjects = 50 women; factor = tube type) showed significant differences across tube types (F(2, 98) = 9.142, p = 0.00023). Post-hoc paired comparisons showed that EDTA tubes yielded significantly higher cfDNA concentrations than both Norgen (p = 0.00042) and Roche tubes (p = 0.02885). Significant differences were also observed between Norgen and Roche tubes (p = 0.01318).

Additional analysis restricted to cfDNA fragments within the 100–250 bp range confirmed a significant effect of blood-collection tube type on cfDNA yield (**Additional File 3**). Repeated measures ANOVA revealed significant differences across tube types (F(2, 98) = 5.003, p = 0.0058). Post-hoc paired comparisons showed significantly higher cfDNA concentrations in EDTA tubes compared to Norgen tubes (p = 0.00548). Significant differences were also observed between Norgen and Roche tubes (p = 0.0193), whereas no significant differences were detected between EDTA and Roche tubes (p = 0.216).

To evaluate the effect of processing delay, samples were stratified according to plasma isolation time (<24 h *versus* >24 h after blood collection). No statistically significant differences in cfDNA concentration were observed between processing-time groups for any blood-collection tube (**Figure 2C**).

Visual inspection revealed varying degrees of hemolysis across samples (**Additional File 4 and Additional File 5**) Norgen tubes showed the highest proportion of hemolyzed plasma samples (24/50, 48%) followed by Roche tubes (21/50, 42%), whereas EDTA tubes were generally minimally hemolyzed (5/50, 10%). Hemolysis was not significantly associated with cfDNA concentration in any tube type (EDTA: p = 0.59; Norgen: p = 0.07; Roche: p = 0.77) (**Additional File 6**).

Collectively, these indicate that blood-collection tube chemistry substantially influences cfDNA recovery in maternal plasma samples, while processing delay and hemolysis do not. Although EDTA tubes yielded higher cfDNA concentrations, the increase likely reflects reduced preservation of blood cells and contamination with genomic DNA released during cell lysis rather than improved recovery of authentic circulating cfDNA, consistent with previous reports (Fernando et al., 2012; Ward Gahlawat et al., 2019).

### Roche tubes improve cfRNA preservation under routine clinical processing conditions

Analysis of cfRNA extracted from maternal blood revealed clear differences in both RNA concentration and quality among the three blood-collection tubes. LabChip electrophoretic profiles showed that all tubes produced the ∼120 nucleotides (nt) 5S ribosomal RNA (rRNA) bands (**Figure 3A, Additional File 2**) and peaks (**Figure 3B**), characteristic of circulating nucleic acids. In addition to the 5S rRNA bands, several additional bands were observed across samples. These bands corresponded to heterogeneous cfRNA fragments and potential background RNA contamination, with variable intensity depending on the sample.

**Figure 3.**
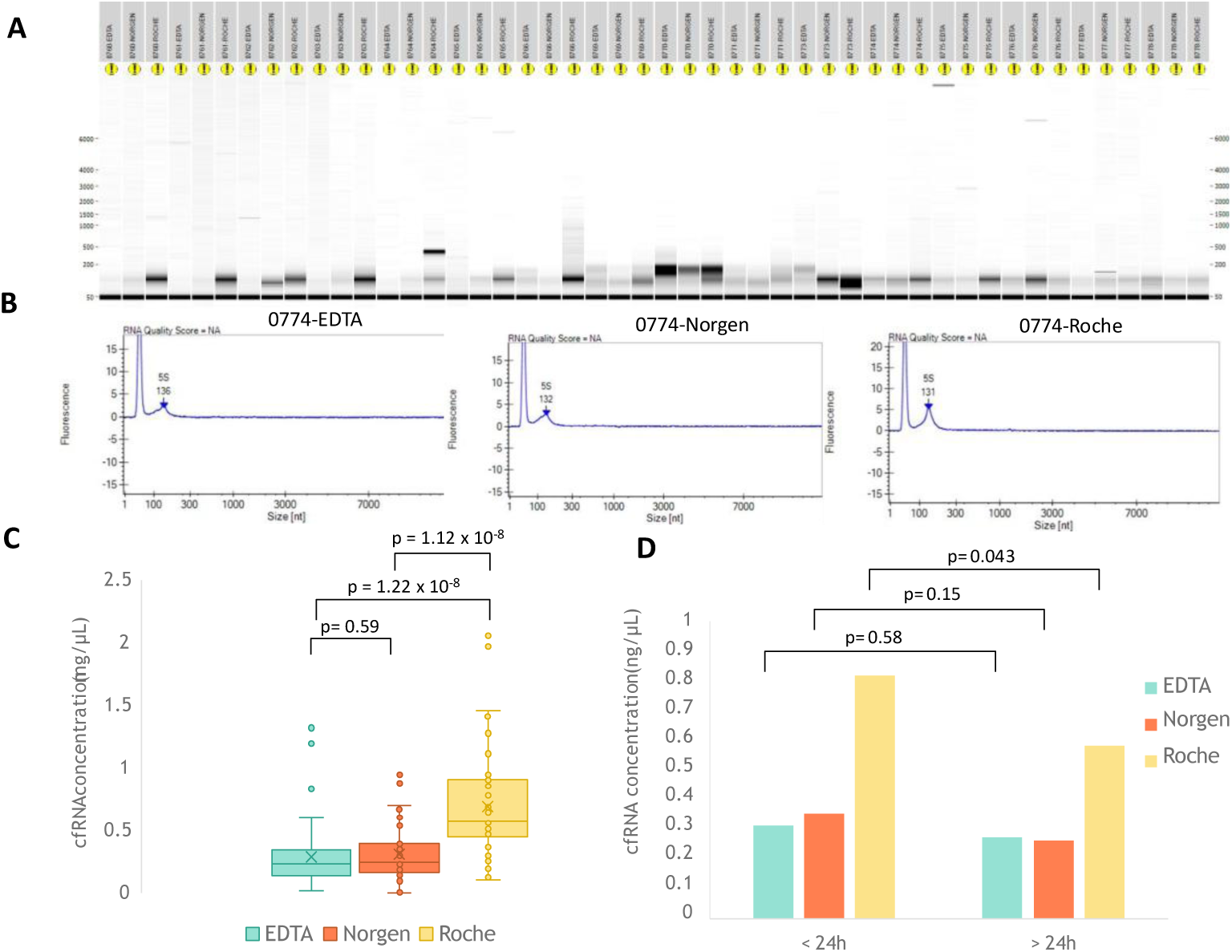
Analysis of cfRNA yield and quality across blood-collection tubes. **A.** Representative electrophoretic profiles obtained by Labchip analysis showing cfRNA fragment size distribution for samples 0760 to 0778 across thethree blood-collectiontubes (EDTA, Norgen and Roche). A clearly defined 5S rRNA peak was observed in 12 out 16 of Roche samples (75%), compared with 4 out of 16 in Norgen samples (25%) and 1 out of 16 in EDTA samples (6.25%). **B.** Representative electropherograms from donor 0774 illustrating cfRNA quality. The first peak belongs to a lower marker, present in every sample. All three tubes display a detectable 5S rRNA peak, typical of plasma-derived cfRNA. **C.** cfRNA concentration (ng/µL) obtained from each tube. A repeated-measures ANOVA (subjects = 50 women; factor = tube type) showed a significant effect of tube type on cfRNA yield (F( 2, 98) = 40.67, p = 1.375 x 10^-13^). Post hoc paired comparisons revealed no significant difference between EDTA and Norgen (p = 0.59), whereas Roche yielded significantly higher cfRNA concentrations than both EDTA (p = 1.22 x 10 ^-8^) and Norgen tubes (p = 1.12 x 10^-8^). **D.** cfRNA concentration (ng/µL) stratified by processing time (< 24 h vs. > 24 h) and grouped by tube type. Post-hoc tests showed no differences between EDTA and Norgen between time-points (p = 0.58 and p = 0.15, respectively), while Roche tubes yielded more cfRNA concentration when processed less than 24 h after sample collection (p = 0.043).

Across the 150 analyzed samples, a clearly defined 5S rRNA peak was observed in 45 of 50 Roche samples (90%), compared with 18 of 50 Norgen samples (36%) and 19 of 50 EDTA samples (38%). Overall, Roche tubes preserved the integrity of cfRNA fragments more consistently, whereas EDTA and Norgen tubes showed greater variability in 5S peak definition.

Quantification of cfRNA concentration further demonstrated a significant effect of blood-collection tube type on cfRNA recovery (**Figure 3C**). A two-way repeated-measures ANOVA confirmed significant differences among tube types (F(2, 98) = 40.67, p = 1.375 × 10^-13^). Post-hoc paired comparisons showed that Roche tubes (0.683 ± 0.417 ng/μL) yielded significantly higher cfRNA concentrations than EDTA tubes (0.289 ± 0.250 ng/μL) (p = 1.22 × 10^-8^) and Norgen tubes (0.305 ± 0.220 ng/μL) (p = 1.12 × 10^-8^), whereas no significant differences were detected between EDTA and Norgen tubes (p = 0.59). These results show that Roche tubes outperform the other two tube types in terms of cfRNA recovery.

As observed for cfDNA, hemolysis was not significantly associated with cfRNA concentration in any tube type (EDTA: p = 0.67; Norgen: p = 0.65; Roche: p = 0.91) (**Additional File 6**), indicating that hemolysis did not substantially affect cfRNA yield in this dataset.

To evaluate whether processing delay influenced cfRNA preservation, samples were stratified according to plasma isolation time (<24 h *versus* >24 h after blood collection) (**Figure 3D**). No significant differences were observed between processing-time groups for EDTA (p = 0.58) or Norgen tubes (p = 0.15). In contrast, Roche tubes yielded significantly higher cfRNA concentrations when processed within 24 h compared to samples processed after 24 h (p = 0.043).

Overall, these results demonstrate that blood-collection tube chemistry substantially influences cfRNA recovery and preservation in maternal plasma samples. Roche tubes consistently yielded higher cfRNA concentrations and more consistent preservation of the characteristic 5S rRNA peak than either EDTA or Norgen tubes. Although delayed processing reduced cfRNA concentration in Roche tubes, their overall performance remained superior. Finally, hemolysis was not significantly associated with cfRNA yield in any tube type.

### Improved cfRNA preservation enhances the detection of subtle biological associations in maternal plasma

To evaluate whether cfRNA concentration reflected transcript abundance in maternal plasma, we examined the relationship between cfRNA concentration and the average Ct value of the housekeeping gene *RPLP0*. As expected, higher cfRNA concentrations correlated with lower Ct values, indicating higher *RPLP0* transcript abundance (**Figure 4**). Linear regression analysis showed a modest inverse association (y = -0.05x + 1.9427, R² = 0.2151), and Pearson correlation analysis confirmed a significant negative correlation between cfRNA concentration and *RPLP0* Ct values (r = -0.4638, p < 1 × 10^-6^). These findings indicate that samples with lower *RPLP0* Ct values yielded proportionally higher cfRNA concentrations and support the use of *RPLP0* as a representative endogenous control for input RNA.

**Figure 4.**
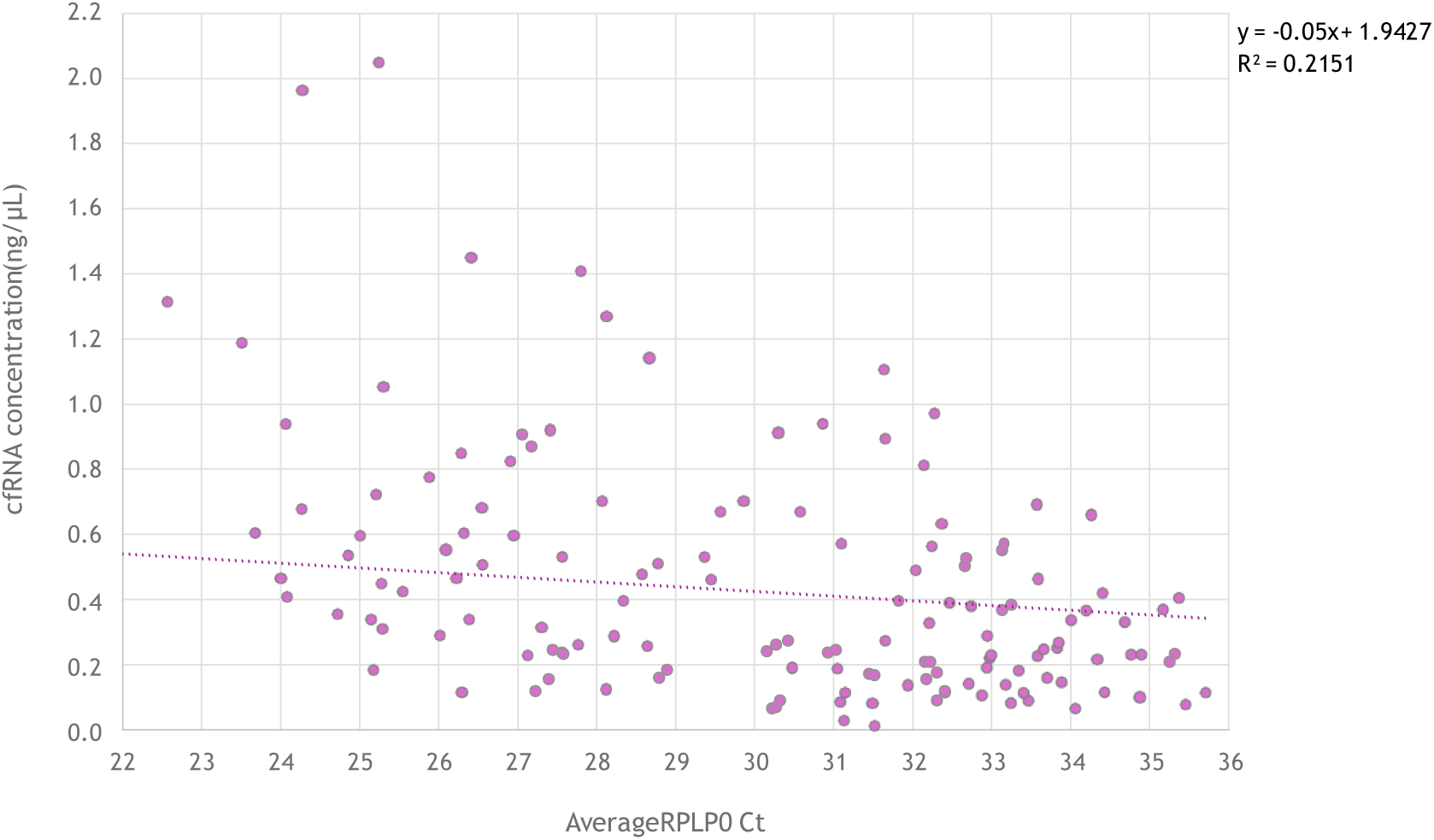
Linear relationshipbetweencfRNA concentrationandthe average *RPLP0* Ct values. Scatter plot showing the linear regression between cfRNA concentration (ng/µL) and the average Ct value of the housekeeping gene *RPLP0*. The association reveals a modest negative correlation, described by the equation y = -0.05x + 1.9427 with an R^2^ = 0.2151. The Pearson correlation coefficient is r = -0.46382714 (p < 1 x 10^-6^).

We next examined whether average *RPLP0* Ct values varied according to GA at delivery. When all samples were analyzed together, a modest correlation between *RPLP0* Ct and GA was observed (p = 0.0303) (**Additional File 7**). After stratification according to GA at birth, a moderate positive correlation was detected in preterm pregnancies (GA < 266 days) (**Figure 5A**). Lower Ct values, reflecting higher transcript abundance, were associated with lower GA before week 38 (r = 0.4743, p = 0.0096). Stratification by tube type revealed similar trends across EDTA (r = 0.4340, p = 0.12), Norgen (r = 0.5908, p = 0.08) and Roche tubes (r = 0.5738, p = 0.06), although none reached statistical significance individually (**Figure 5B**). In contrast, no meaningful associations were observed in term pregnancies (GA > 266 days), either when all tubes were analyzed together (r = -0.0601, p = 0.2555) (**Figure 5C**) or after tube-specific stratification (EDTA: r = -0.0362, p = 0.41; Norgen: r = -0.0760, p = 0.31; Roche: r = -0.1021, p = 0.25) (**Figure 5D**).

**Figure 5.**
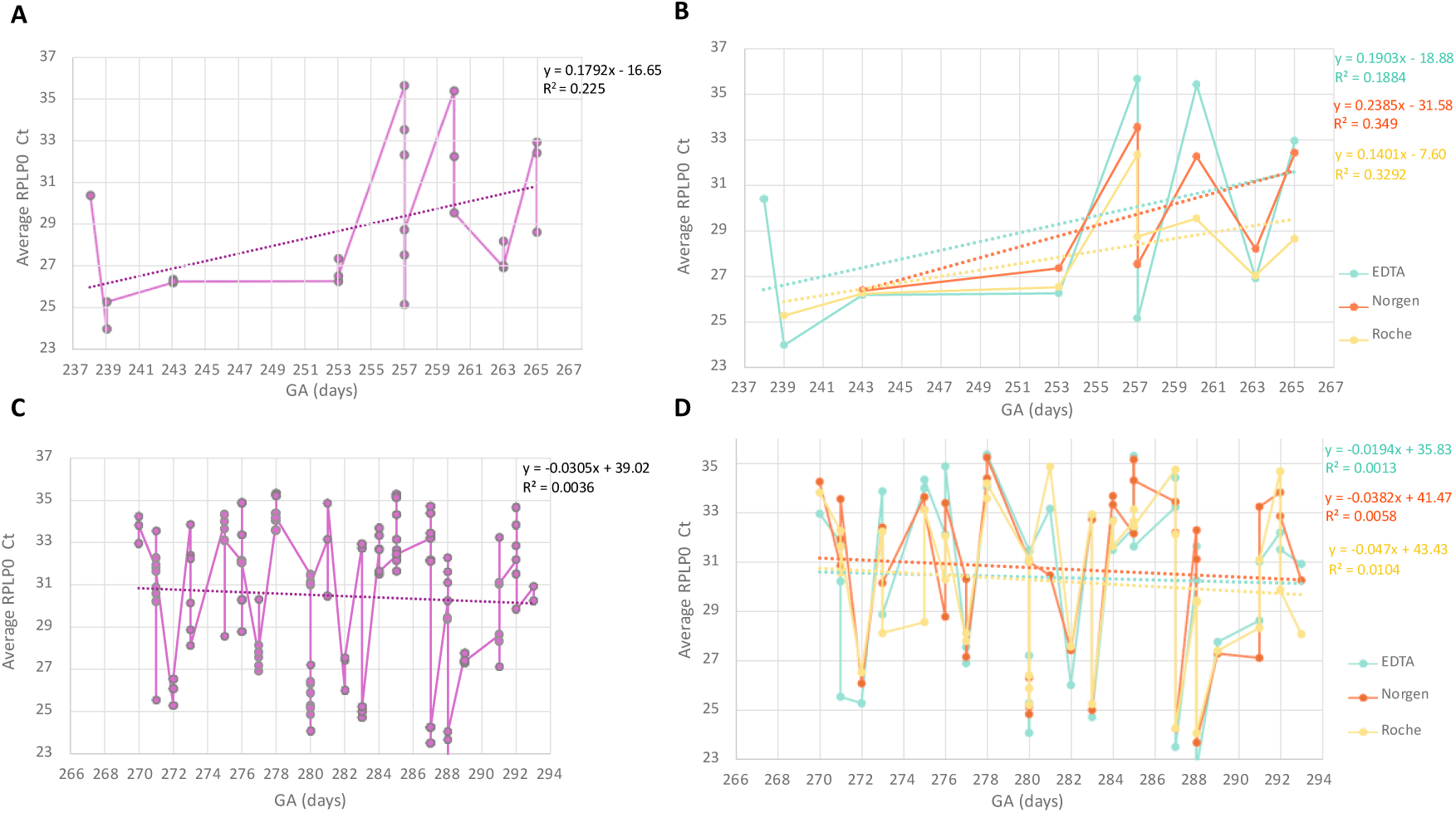
Linear relationship between average *RPLP0* Ct values and GA in preterm and term pregnancies. **A.** Linear regression between the average *RPLP0* Ct values and preterm samples (GA < 266 days) gestation days with all tubes combined. A positive correlation is observed (r = 0.4743, p = 0.0096). **B.** Same analysisrestricted to preterm samplesbut stratified by tube type. Pearson’s correlation coefficients: EDTA r = 0.4340 (p = 0.12), Norgen r = 0.5908 (p = 0.08), and Roche r = 0.5738 (p = 0.06). **C.** Linear regression between the average *RPLP0* Ct values and term samples (GA > 266 days) gestation days with all tubes combined. No meaningful association is observed (r = -0.0601, p = 0.2555). **D.** Same analysis restricted to term samples but stratified by tube type. Pearson’s correlation coefficients: EDTA r = -0.0362 (p = 0.41), Norgen r = -0.0760 (p = 0.31), and Roche r = -0.1021 (p = 0.25).

We subsequently evaluated the relative expression of *GDF15* and *ARHGEF28* in maternal plasma. *ARHGEF28* results were excluded from further analyses because the gene showed high Ct values (>35 mean Ct per sample), poor reproducibility between replicates and irregular melting-curve profiles, including variable peak shapes and absence of defined melting peaks in several samples (**Additional File 8**). These findings indicated unstable amplification and prevented reliable quantification under the experimental conditions used.

Analysis of *GDF15* relative expression revealed no significant association with GA when all samples were analyzed together (r = 0.0481, p = 0.2834) (**Additional File 9**). However, after stratification by tube type, moderate positive correlations between *GDF15* expression and GA were observed in both Norgen (r = 0.2161, p = 0.00395) and Roche tubes (r = 0.2153, p = 0.0041), whereas EDTA samples showed a non-significant negative trend (r = -0.088, p = 0.14) (**Figure 6**).

**Figure 6.**
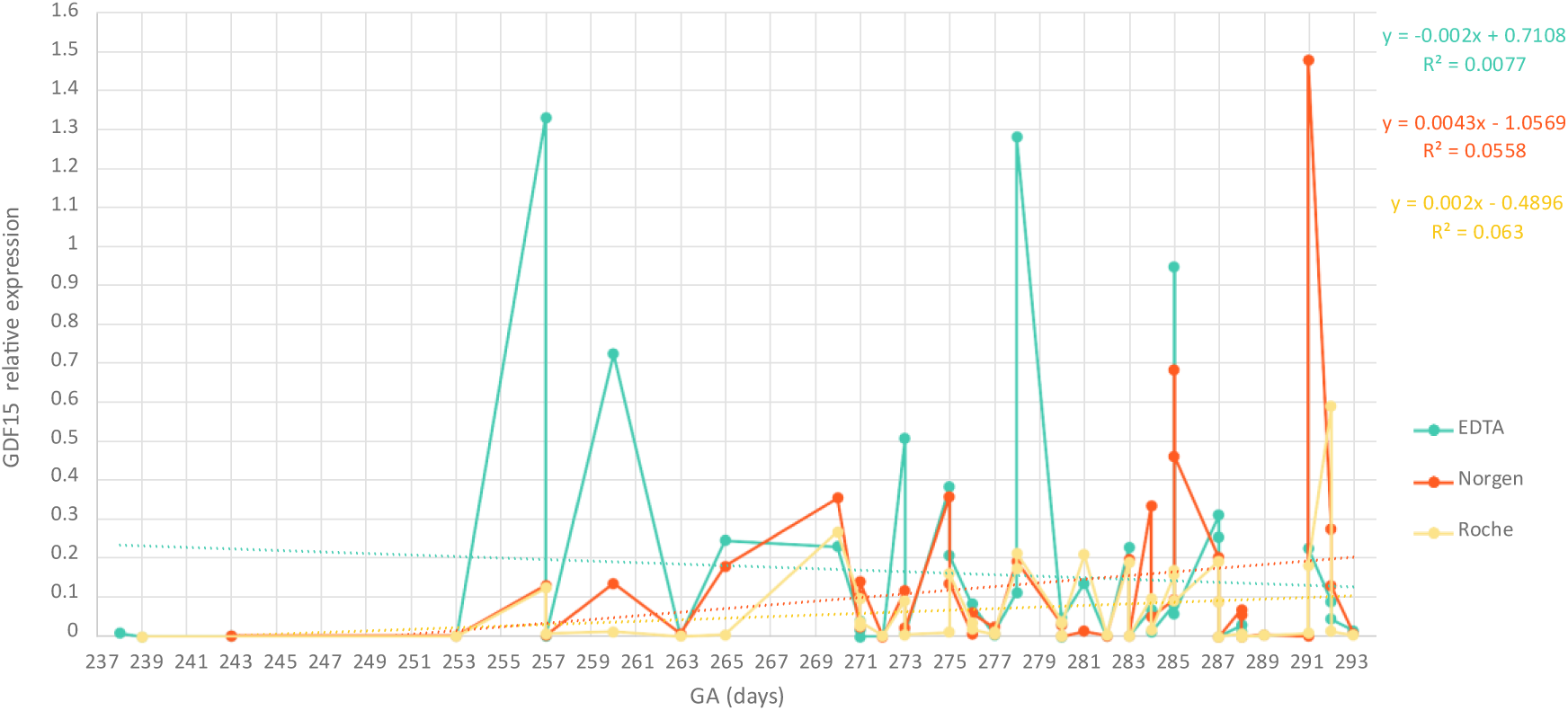
Linear relationship between *GDF15* relative expression and GA stratified by tube type. Scatter plot showing the linear regression between the relative expression of *GDF15* and GA (days) across different tube types. Pearson’s correlation coefficients revealedstatisticallysignificant positive associations for Norgen (r = 0.2161, p = 0.00395) and Roche (r = 0.2153, p = 0.0041), whereas EDTA showed no significant correlation (r = -0.088, p = 0.14).

We next evaluated the effect of processing delay on *GDF15* expression. No significant differences in *GDF15* relative expression were observed when samples were grouped according to processing time alone (<24 h versus >24 h) (p = 0.17) **(Figure 7A**). After stratification by tube type, no significant differences associated with processing delay were detected in EDTA (p = 0.53) or Norgen tubes (p = 0.66), whereas Roche tubes showed a modest but statistically significant increase in *GDF15* expression in samples processed after 24 h (p = 0.013) (**Figure 7B**).

**Figure 7.**
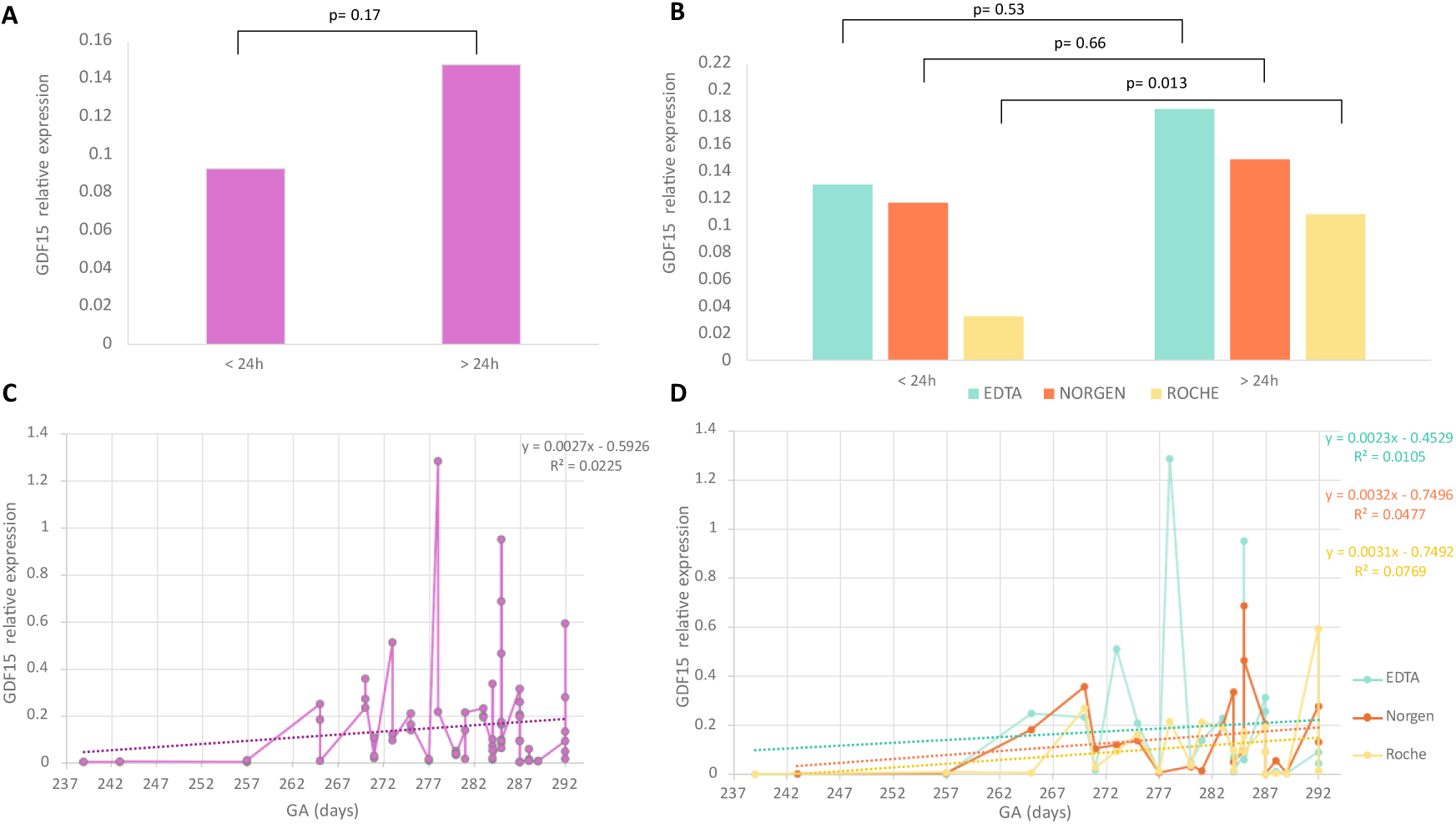
Effect of processing time on *GDF15* relative expression. **A.** *GDF15* relative expression as a function of processingtime, comparingsamplesprocessedin < 24 h versus > 24 h. No significant difference was observed (p = 0.17). **B.** Same comparison stratified by tube type. Pairwise comparisons show no differences between EDTA (p = 0.53) and Norgen (p = 0.66) regardless of processing time, while Roche shows a modest but significant difference (p = 0.013). **C.** Linear regression between *GDF15* relative expression and GA with all the tubes combined in samples processed more than 24 h after collection. A weak, non-significant positive correlation was observed (r = 0.15, p = 0.0994). **D.** Same analysis stratified by tube type. Pearson’s correlation coefficients were calculated: EDTA r = 0.1025 (p = 0.3091), Norgen r = 0.2184 (p = 0.1527) and Roche r = 0.2773 (p = 0.0898). Across all analyses, no statistically significant associations were detected between *GDF15* and GA in samples processed more than 24 hours after collection.

Finally, we examined whether *GDF15* expression correlated with GA according to processing time. No significant associations were detected in samples processed with in 24 h (**Additional File 10**). In samples processed after 24 h, a weak non -significant positive correlation was observed between *GDF15* expression and GA when all tube types were analyzed together (r = 0.15, p = 0.0994) (**Figure 7C**). Similar non-significant trends were observed after tube-specific stratification (EDTA: r = 0.1025, p = 0.3091; Norgen: r = 0.2184, p = 0.1527; Roche: r = 0.2773, p = 0.0898) (**Figure 7D**).

In summary, these findings suggest that preanalytical conditions influence not only cfRNA recovery, but also the sensitivity to detect subtle biological associations in maternal plasma. While *RPLP0* Ct values showed GA-dependent patterns in preterm pregnancies, *GDF15* expression displayed modest tube-dependent associations with GA that were more evident in preservative-containing tubes.

## Discussion

In the present study, we describe the implementation of the Registry of Pregnant Women at Cruces University Hospital, a prospective pregnancy registry and biobanking infrastructure established at a tertiary-care academic hospital in the Basque Country. We also report a preanalytical substudy evaluating the impact of blood-collection tube type and processing delay on circulating nucleic acid preservation in maternal plasma collected at delivery. Within the context of increasingly evolving DOHaD-oriented and translational pregnancy research, the establishment of standardized infrastructures enabling systematic collection of maternal plasma and placental samples, under ethically controlled conditions, constitutes a critical step toward future clinically integrated molecular studies. In this context, our findings indicate that preanalytical variables substantially influence the recovery, integrity and biological interpretability of circulating nucleic acids, particularly cfRNA, under routine clinical conditions.

Regarding cfDNA, electrophoretic profiles showed the characteristic fragmentation pattern characterized by a dominant mono-nucleosomal peak around 160–180 bp together with additional higher-order fragments, consistent with previous descriptions of circulating cfDNA^14^. Quantitative analyses demonstrated that EDTA tubes yielded higher cfDNA concentrations than both Norgen and Roche tubes. However, previous studies suggest that increased cfDNA concentration in EDTA tubes likely reflects contamination with genomic DNA released during *ex vivo* blood-cell lysis rather than improved preservation of authentic circulating cfDNA^11,15^. In contrast, preservative-containing tubes are specifically designed to stabilize blood cells and minimize cellular degradation, thereby reducing genomic DNA contamination. Our observations highlight the importance of blood-collection tube chemistry when interpreting cfDNA measurements in maternal plasma studies.

The most pronounced differences across blood-collection tubes were observed for cfRNA preservation. Roche tubes consistently outperformed EDTA and Norgen tubes in both cfRNA concentration and electrophoretic quality. Although all tube types preserved the characteristic ∼120 nt 5S rRNA peak typical of circulating nucleic acids^16^, Roche tubes showed substantially cleaner electrophoretic profiles and more consistent preservation of this signal across samples. In contrast, EDTA and Norgen tubes displayed greater variability together with additional heterogeneous bands compatible with fragmented RNA populations and potential background contamination^17^. Quantitative analyses further confirmed significantly higher cfRNA recovery in Roche tubes. Collectively, our results identify tube chemistry as a major determinant of cfRNA preservation and downstream transcript detectability in maternal plasma.

Our results also highlight the importance of standardized processing workflows within prospective pregnancy registries and biobanking infrastructures. Although Roche tubes maintained superior overall performance under delayed processing conditions, cfRNA concentrations significantly decreased when plasma isolation occurred after more than 24 hours. These observations reinforce the importance of minimizing processing delays whenever possible, even when preservative-containing tubes are used. Importantly, one of the central objectives of the registry described here was precisely the establishment of harmonized and reproducible workflows compatible with routine clinical activity at delivery rooms and future longitudinal molecular analyses.

Another important observation relates to hemolysis. Although Norgen and Roche tubes showed higher frequencies of visible hemolysis than EDTA tubes, hemolysis was not significantly associated with cfDNA or cfRNA yield in any tube type. These findings suggest that the differences observed across tubes are primarily driven by preservation chemistry rather than by hemolysis itself, consistent with previous reports^9^. Notably, Roche tubes maintained superior cfRNA recovery and cleaner electrophoretic profiles despite relatively frequent hemolysis, further supporting their robustness for maternal plasma cfRNA studies.

Beyond technical performance, our findings suggest that improved cfRNA preservation may enhance the sensitivity to detect subtle biological variability in maternal plasma. The inverse correlation observed between cfRNA concentration and *RPLP0* Ct values supports that higher cfRNA yields correspond to higher endogenous transcript abundance and supports the reliability of the extraction and quantification workflow used in this study. Notably, *RPLP0* Ct values showed GA-dependent patterns specifically in pregnancies delivering before 38 weeks of gestation, whereas no meaningful association was observed in term pregnancies. Although the number of preterm pregnancies included in the substudy was limited, these observations suggest that earlier deliveries may be characterized by increased circulating cfRNA abundance that stabilizes near term. Previous studies have associated earlier delivery with placental stress, oxidative stress and inflammatory activation^18,19^, processes that could contribute to altered circulating nucleic acid profiles.

Similarly, *GDF15* expression showed modest positive associations with GA that were detectable mainly in preservative-containing tubes. *GDF15* is a stress-responsive cytokine associated with placental stress, inflammation and adverse pregnancy outcomes^20,21^. Previous studies have linked elevated *GDF15* levels to preterm birth and preeclampsia, particularly under conditions of increased placental stress^20,21^. However, we did not observe evidence of markedly increased *GDF15* expression in pregnancies delivering at earlier GAs, likely because the study population mainly consisted of term pregnancies and did not include very early or extreme preterm births. Although the present study was not designed for biomarker discovery or clinical prediction, the observation that subtle associations with GA were more consistently detectable in Roche and Norgen tubes is consistent with the hypothesis that improved preanalytical preservation may increase sensitivity for detecting biologically meaningful variability in maternal plasma cfRNA. In contrast, *ARHGEF28* amplification was not sufficiently robust under the present experimental conditions, highlighting the technical challenges associated with low-abundance circulating transcripts.

Interestingly, delayed processing modestly increased *GDF15* expression in Roche tubes despite the overall reduction in cfRNA concentration observed after prolonged storage. One possible explanation is that stress-responsive transcripts may be particularly sensitive to *ex vivo* cellular activation during delayed sample handling. Previous studies have shown that prolonged storage conditions may alter transcriptional profiles through hypoxia-related and cellular stress pathways^10,22^. These observations further emphasize that preanalytical handling may influence not only RNA integrity, but also the apparent biological interpretation of circulating transcriptomic data.

The present study should also be interpreted within the context of its ethical and infrastructural framework. One of the defining characteristics of the Registry of Pregnant Women at Cruces University Hospital is that future access to maternal and offspring clinical records requires independent ethics approval and explicit participant consent, ensuring strict compliance with data-protection and governance standards. Although this model limits the immediate availability of extensive clinical phenotyping, it establishes a robust ethical foundation for future longitudinal and clinically integrated studies, and enables the systematic collection of biological samples in clinical routine. In this context, the implementation of reliable and standardized preanalytical workflows becomes particularly relevant, as the long-term value of prospective registries ultimately depends on the quality, reproducibility and interpretability of stored biological material.

Several limitations should be acknowledged. First, the sample size of the preanalytical substudy was relatively small and included only a limited number of pregnancies delivering before 38 weeks of gestation, reducing statistical power for GA-related analyses. Second, the study was based exclusively on samples collected at delivery and therefore does not address longitudinal changes during pregnancy. Third, the analyses focused on a limited number of transcripts and were not designed for biomarker discovery or clinical prediction. Despite these limitations, the study provides a detailed characterization of preanalytical variability under real-world clinical conditions and establishes a standardized workflow integrated within a prospective pregnancy registry and biobanking infrastructure.

## Conclusion

The present work clearly shows that preanalytical conditions critically influence cfRNA preservation, transcript detectability and the ability to identify subtle biological associations in maternal plasma. Although EDTA tubes yielded higher cfDNA concentrations, these increases likely reflect genomic DNA contamination resulting from blood-cell lysis rather than improved preservation of circulating nucleic acids. In contrast, Roche tubes provided cleaner electrophoretic profiles, improved cfRNA recovery and more consistent detection of biologically meaningful associations, although delayed processing reduced this advantage. More broadly, our results highlight the importance of integrating rigorous preanalytical standardization into prospective pregnancy registries and translational biobanking initiatives aimed at future molecular studies of maternal, placental and offspring health.

## Data Availability

Data generated from the Registry will be made publicly available when generated and published. Samples can be requested upon approval of the local Ethics Board, and are subjected to availability in the Basque Biobank.

## Additional files

**Additional File 1: Summary table of donor information.** Donor information including donor code and GA expressed in completed weeks, days, and total days. Note: The donor codes listed in this dataset are pseudonymized research identifiers generated during the biobank registration process. These codes are strictly confidential and secure so cannot be linked to any personal identifiable information by any unauthorized person or anyone outside the research group. (.xlsx 11 KB)

**Additional File 2: Electrophoretic profiles showing fragment distribution across blood - collection tubes.** A. Electrophoreic profiles obtained by Labchip analysis showing cfDNA fragment size distribution for samples across the three blood-collection tubes (EDTA, Norgen and Roche). B. Electrophoretic profiles obtained by Labchip analysis showing cfRNA fragment size distribution for samples 0760 to 0778 across the three blood-collection tubes (EDTA, Norgen and Roche). (.pdf 346 KB)

**Additional File 3: Analysis of cfDNA yield and fragment distribution across blood-collection tubes.** cfDNA concentration (ng/µL) quantified within the 100-250 bp range. A repeated measures ANOVA (subjects = 50 women; factor = tube type) revealed a significant effect of tube type on cfDNA yield (F(2, 98) = 5.003, p = 0.0058). Post-hoc paired comparisons showed that EDTA tubes yielded significantly higher cfDNA concentrations than Norgen (p = 0.00548). Significant differences were also observed between Norgen and Roche (p = 0.0193), but not between EDTA and Roche (p = 0.216). (.pdf 36 KB)

**Additional File 4: Visual assessment of hemolysis across blood-collection tubes.** Representative appearance of plasma obtained from EDTA (left), Roche (middle) and Norgen (right) tubes. EDTA sample showed no visible hemolysis, Roche sample displayed mild hemolysis, and Norgen sample exhibited marked hemolysis. (.pdf 120 KB)

**Additional File 5: Number of hemolyzed samples by tube type.** The table summarizes the frequency of hemolysis observed in plasma samples collected in EDTA, Norgen and Roche tubes. Norgen tubes showed the highest proportion of hemolyzed samples (24/50) (48%), followed by Roche (21/50) (42%), whereas EDTA tubes exhibited minimal hemolysis (5/50) (10%). (.xlsx 12 KB)

**Additional File 6: Effect of hemolysis in cfDNA and cfRNA concentration.** A. cfDNA concentration (ng/µL) stratified by observable hemolysis (hemolysis vs. no hemolysis) in samples and grouped by tube type. *Post hoc* tests showed no differences in cfDNA concentration in either tube, regardless of hemolysis or not (EDTA: p = 0.59, Norgen: p = 0.07 and Roche: p = 0.77). B. cfRNA concentration (ng/µL) stratified by observable hemolysis (hemolysis vs. no hemolysis) and grouped by tube type. *Post hoc* tests showed no differences in cfRNA concentration in either tube, regardless of hemolysis or not (EDTA: p = 0.67, Norgen: p = 0.65 and Roche: p = 0.91). (.pdf 52 KB)

**Additional File 7: Linear relationship between average *RPLP0* Ct values and GA.** Scatter plot showing the linear regression between average *RPLP0* Ct value**s** and GA (days) with all the tubes combined. The association reveals a positive correlation, described by the equation y = 0.0428x + 18.32 with an R^2^ = 0.0241. The Pearson correlation coefficient is r = 0.1552 (p = 0.0303), indicating a modest statistical significance between average *RPLP0* Ct values and GA of birth. (.pdf 41 KB)

**Additional File 8: Melt curves and melt peak analysis of the *ARHGEF28* RT-qPCR assay.** A. Melt curve profiles showing fluorescence decay across increasing temperatures, revealing inconsistent amplification patterns. B. Derivative melt peaks displaying variable shapes, and, in several cases, the absence of a defined melting peak. (.pdf 207 KB)

**Additional File 9: Linear relationship between *GDF15* relative expression and GA.** Scatter plot showing the linear regression between the relative expression of *GDF15* and GA (days). The association reveals a modest positive correlation, described by the equation y = 0.001x – 0.1428 with an R^2^ = 0.0023. The Pearson correlation coefficient is r = 0.0481 (p = 0.2834), indicating a non-significant relationship between the *GDF15* relative expression and the GA of birth. (.pdf 41 KB)

**Additional File 10: Effect of processing time on *GDF15* relative expression in samples processed within 24 h after collection.** A. Linear regression between *GDF15* relative expression and GA with all the tubes combined in samples processed within 24 h after collection. A weak, non-significant negative correlation was observed (r = -0.0757, p = 0.2680). B. Same analysis stratified by tube type. Pearson’s correlation coefficients were calculated: EDTA r = -0.3255 (p = 0.0648), Norgen r = 0.2229 (p = 0.1532) and Roche r = -0.0193 (p = 0.4652). Across all analyses, no statistically significant associations were detected between *GDF15* and GA in samples processed within 24 hours after collection. (.pdf 94 KB)

## Declarations

### Ethics approval and consent to participate

The Registry of Pregnant Women at Cruces University Hospital was approved by the Research Ethics Committee of the Integrated Health Organisation Ezkerraldea-Enkarterri-Cruces (Comité de Ética de la Investigación OSI Ezkerraldea-Enkarterri-Cruces; approval no. CEI E23-50). The present preanalytical substudy received independent approval from the same committee (approval no. CEI E25-46).

### Authors’ contributions

I.G.-M., H.S.-G., J.R.B., J.B. and N.F.-J. conceived and designed the study. T.M.C., M.P.E.L., E.Q.O., P.O.S., J.O.C., A.A.A., C.O.S., M.M.G., L.R.G., M.J.M.M. and J.B. supervised the clinical procedures and the sample collection. A.R.L., S.E.G., L.M.C. and R.C.G. designed the sample processing workflow and helped with the ethical aspects of the project. A.S.P., A.R.A., A.M.I., A.A.O., A.A.G., A.G.G., A.S.S., A.H.C., A.F.S., A.I.R.J., A.I.T.L., A.J.C.O., A.M.M.T., A.S.H., A.S.D., A.B.M., A.C.L., A.E.V., B.G.D., B.L.M., B.D.A., C.M.L., C.G.R., C.N.P., C.G.T., C.B.C., E.C.H., E.A.A., E.M.V., E.G.A.A., G.M.C., I.Z.P., I.H.O., I.P.P., I.S.M., I.C.Q., I.R.P., I.R.C., J.G.C., J.D.I., J.S.M., L.M.A., L.A.P., L.V.R.L., L.O.S., L.S.C., L.V.G., M.A.B., M.T.S., M.C.M., M.F.A., M.V.B., M.A.L., M.C.M., M.M.I., M.B. G., M.I.R., M.S.V., N.L.Q., N.M.P., N.O.C., N.A.R., N.E.A., N.M.T., N.A.G., N.T.L., O.P.R., P.S.Z., P.G.O., P.O.S., R.L.R., R.S.A., S.G.S., S.M.M., S.M.M., S.M.F., S.R.M., S.U.M., S.A.B., S.T.E., T.M.U., V.G.S., V.V.A., Y.V.I., Y.M.I., Z.Z.I., Z.C.R. explained the informed consent to the mothers and the families, underwent specific formation on sample handling, advised to improve the protocols and collected the samples of both the Registry and the substudy. I.G.-M., H.S.-G., M.P.-S., N.S.-B. and I.G.-S. performed the experiments of the substudy and analyzed the data. I.G.-M., H.S.-G., J.R.B. and N.F.-J. drafted the manuscript. J.B. and N.F.-J. jointly directed the study. All authors revised and approved the final manuscript.

## Acknowledgements

We would like to thank the mothers, children and families participating in the Registry of Pregnant Women at Cruces University Hospital, as well as the orderlies who transported the samples daily to the Basque Biobank. The authors also thank for the technical and human support provided by SGIker (EHU/ERDF, EU). Finally, we would like to thank the Basque Biobank, the Basque Public Health Service (Osakidetza), the management of Cruces University Hospital, and the Department of Health of the Basque Government for the human support and for providing many of the essential supplies required for sample collection. The present project is the result of a large-scale, voluntary, collective and multidisciplinary effort, and is entirely dependent on the universal and public nature of our healthcare system.

## References

1. Jain VG, Monangi N, Zhang G, Muglia LJ. Genetics, epigenetics, and transcriptomics of preterm birth. Am J Reprod Immunol. 2022;88(4):e13600. doi:10.1111/aji.13600

2. Cilleros-Portet A, Lesseur C, Marí S, et al. Potentially causal associations between placental DNA methylation and schizophrenia and other neuropsychiatric disorders. Nat Commun. 2025;16(1):2431. doi:10.1038/s41467-025-57760-3

3. Gray KJ, Hemberg M, Karumanchi SA. Cell-Free RNA Transcriptome and Prediction of Adverse Pregnancy Outcomes. Clin Chem. 2022;68(11):1358–1360. doi:10.1093/clinchem/hvac109

4. Munchel S, Rohrback S, Randise-Hinchliff C, et al. Circulating transcripts in maternal blood reflect a molecular signature of early-onset preeclampsia. Sci Transl Med. 2020;12(550):eaaz0131. doi:10.1126/scitranslmed.aaz0131

5. Camunas-Soler J, Gee EPS, Reddy M, et al. Predictive RNA profiles for early and very early spontaneous preterm birth. American Journal of Obstetrics & Gynecology. 2022;227(1):72.e1-72.e16. doi:10.1016/j.ajog.2022.04.002

6. Krasnyi AM, Sadekova AA, Vtorushina VV, Кan NE, Tyutyunnik VL, Krechetova LV. Extracellular DNA levels and cytokine profiles in preterm birth: a cohort study. Arch Gynecol Obstet. 2022;306(5):1495–1502. doi:10.1007/s00404-022-06456-w

7. Karapetian АО, Baev ОR, Sadekova АА, Krasnyi АМ, Sukhikh GT. Cell-Free Foetal DNA as a Useful Marker for Preeclampsia Prediction. Reprod Sci. 2021;28(5):1563–1569. doi:10.1007/s43032-021-00466-w

8. Jin H, Zhang Y, Fan Z, et al. Identification of novel cell-free RNAs in maternal plasma as preterm biomarkers in combination with placental RNA profiles. J Transl Med. 2023;21:256. doi:10.1186/s12967-023-04083-w

9. Fernando MR, Norton SE, Luna KK, Lechner JM, Qin J. Stabilization of cell-free RNA in blood samples using a new collection device. Clinical Biochemistry. 2012;45(16-17):1497–1502. doi:10.1016/j.clinbiochem.2012.07.090

10. Jiang Z, Lu Y, Shi M, Li H, Duan J, Huang J. Effects of storage temperature, storage time, and hemolysis on the RNA quality of blood specimens: A systematic quantitative assessment. Heliyon. 2023;9(6):e16234. doi:10.1016/j.heliyon.2023.e16234

11. Ward Gahlawat A, Lenhardt J, Witte T, et al. Evaluation of Storage Tubes for Combined Analysis of Circulating Nucleic Acids in Liquid Biopsies. Int J Mol Sci. 2019;20(3):704. doi:10.3390/ijms20030704

12. Hospital Universitario Cruces. https://www.osakidetza.euskadi.eus/osi-ezkerraldea-enkarterri-cruces-hospital-universitario-presentacion/webosk00-ezenccon/es/ Accessed 20 February 2026.

13. Livak KJ, Schmittgen TD. Analysis of relative gene expression data using real-time quantitative PCR and the 2(-Delta Delta C(T)) Method. Methods. 2001;25(4):402–408. doi:10.1006/meth.2001.1262

14. Ungerer V, Bronkhorst AJ, Uhlig C, Holdenrieder S. Cell-Free DNA Fragmentation Patterns in a Cancer Cell Line. Diagnostics (Basel). 2022;12(8):1896. doi:10.3390/diagnostics12081896

15. Andersson D, Kristiansson H, Luna Santamaría M, et al. Evaluation of automatic cell free DNA extraction metrics using different blood collection tubes. Sci Rep. 2025;15(1):19364. doi:10.1038/s41598-025-03508-4

16. Ciganda M, Williams N. Eukaryotic 5S rRNA biogenesis. Wiley Interdiscip Rev RNA. 2011;2(4):523–533. doi:10.1002/wrna.74

17. Zhong P, Bai L, Hong M, et al. A Comprehensive Review on Circulating cfRNA in Plasma: Implications for Disease Diagnosis and Beyond. Diagnostics. 2024;14(10):1045. doi:10.3390/diagnostics14101045

18. Couture C, Brien ME, Boufaied I, et al. Proinflammatory changes in the maternal circulation, maternal–fetal interface, and placental transcriptome in preterm birth. American Journal of Obstetrics & Gynecology. 2023;228(3):332.e1-332.e17. doi:10.1016/j.ajog.2022.08.035

19. Lv M, Jia Y, Dong J, Wu S, Ying H. The landscape of decidual immune cells at the maternal– fetal interface in parturition and preterm birth. Inflamm Res. 2025;74(1):44. doi:10.1007/s00011-025-02015-6

20. Almudares F, Hagan J, Chen X, Devaraj S, Moorthy B, Lingappan K. Growth and differentiation factor 15 (GDF15) levels predict adverse respiratory outcomes in premature neonates. Pediatr Pulmonol. 2023;58(1):271–278. doi:10.1002/ppul.26197

21. Menon R, Richardson LS, Lappas M. Fetal membrane architecture, aging and inflammation in pregnancy and parturition. Placenta. 2019;79:40–45. doi:10.1016/j.placenta.2018.11.003

22. Xing Y, Yang X, Chen H, et al. Impact of storage conditions on peripheral leukocytes transcriptome. Mol Biol Rep. 2021;48(2):1151–1159. doi:10.1007/s11033-021-06194-3

