## Supplementary figures and images for "The Registry of Pregnant Women at Cruces University Hospital: an ethical framework for prospective research with preanalytical optimization of maternal plasma processing"

### Additional File 2

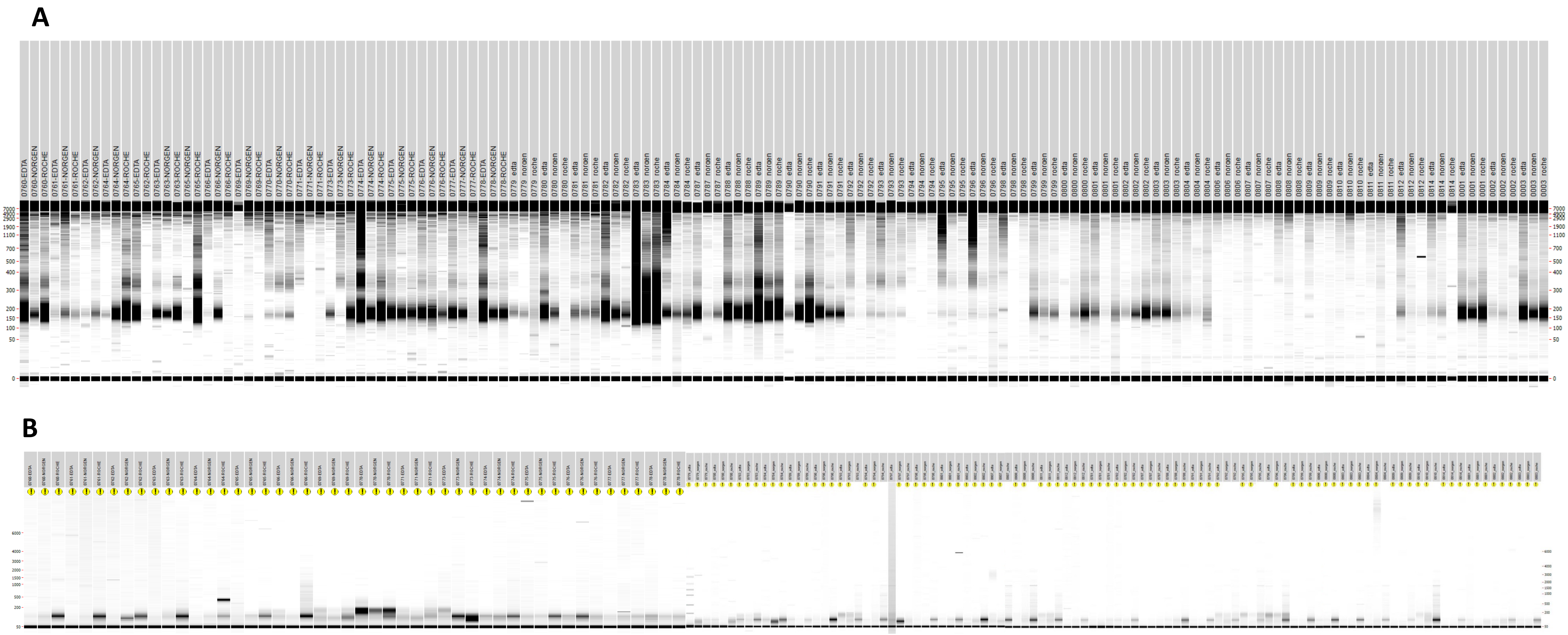

### Additional File 3

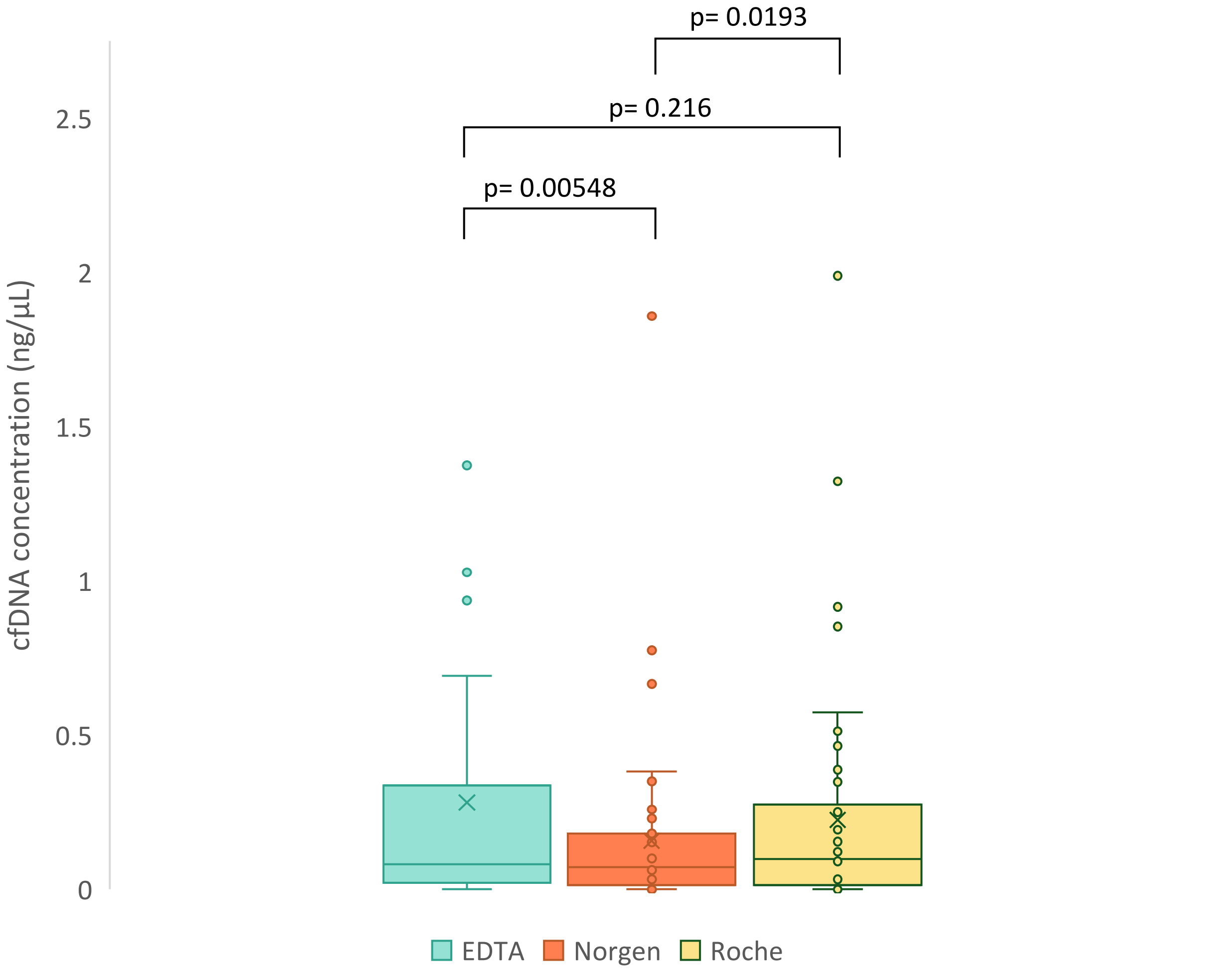

### Additional File 4

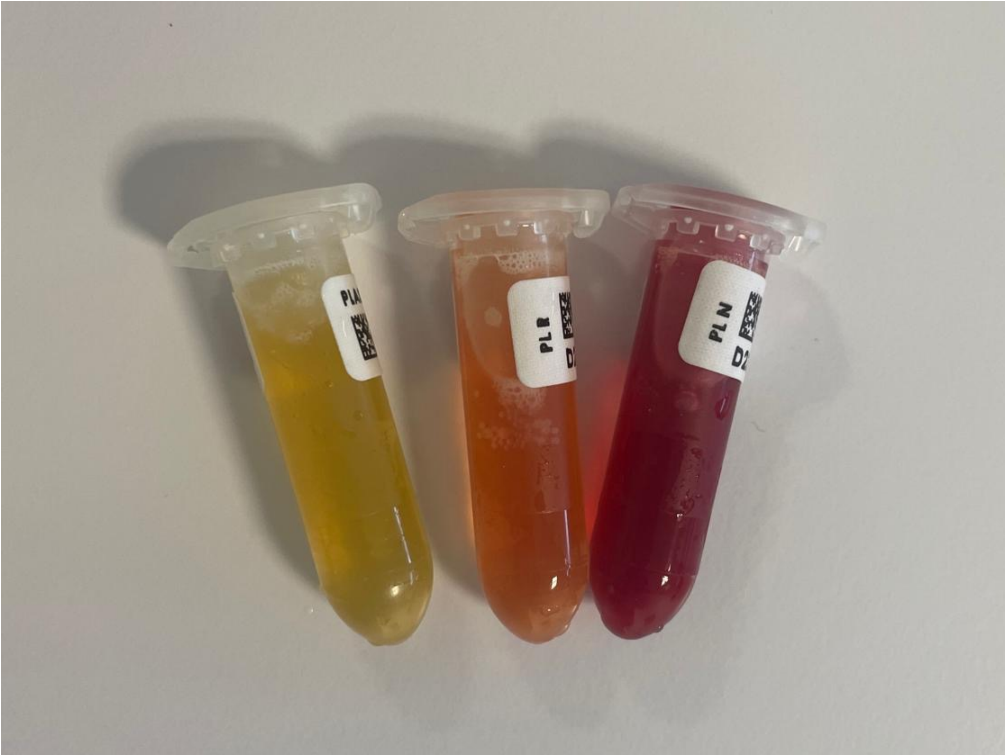

### Additional File 6

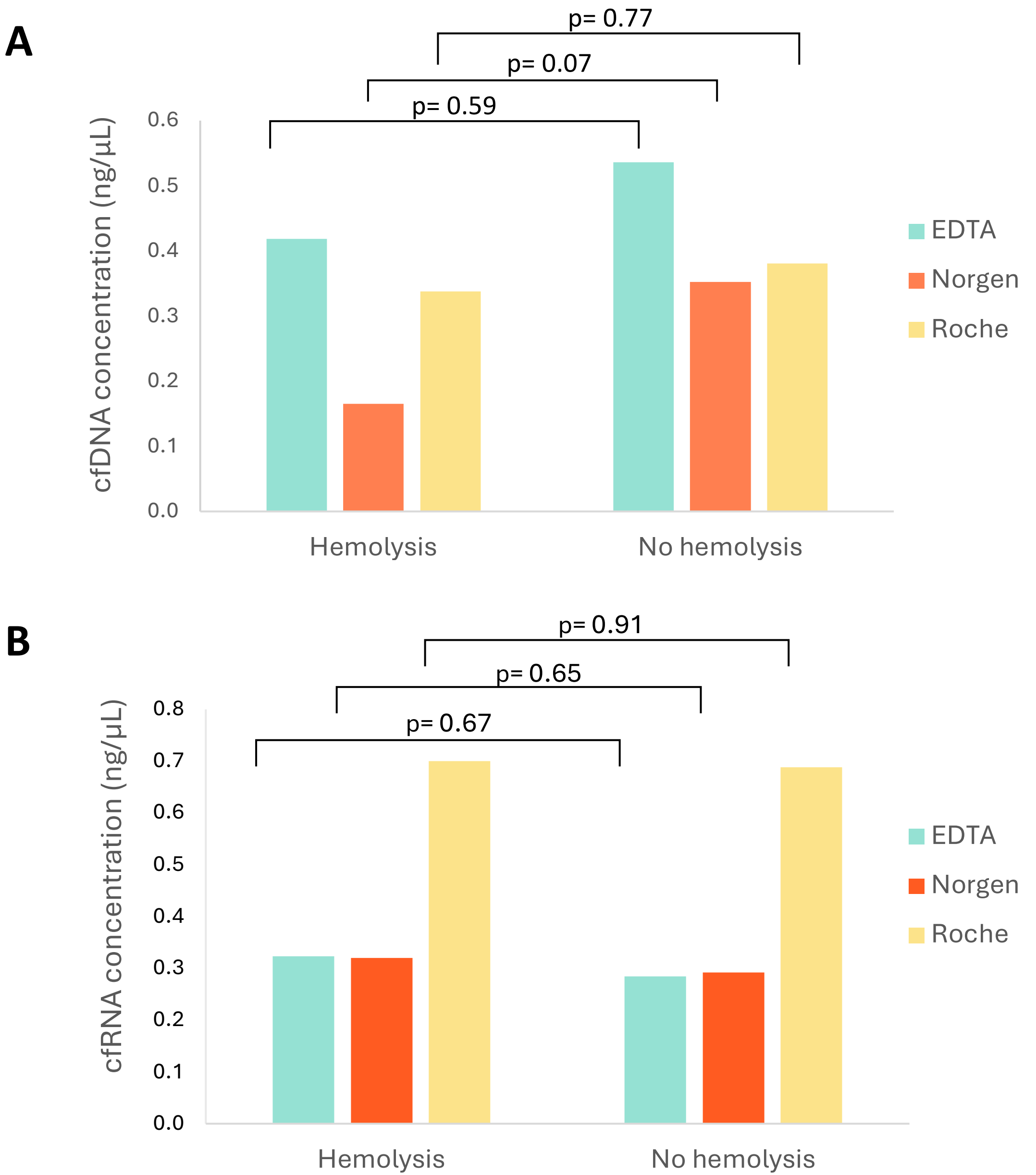

### Additional File 7

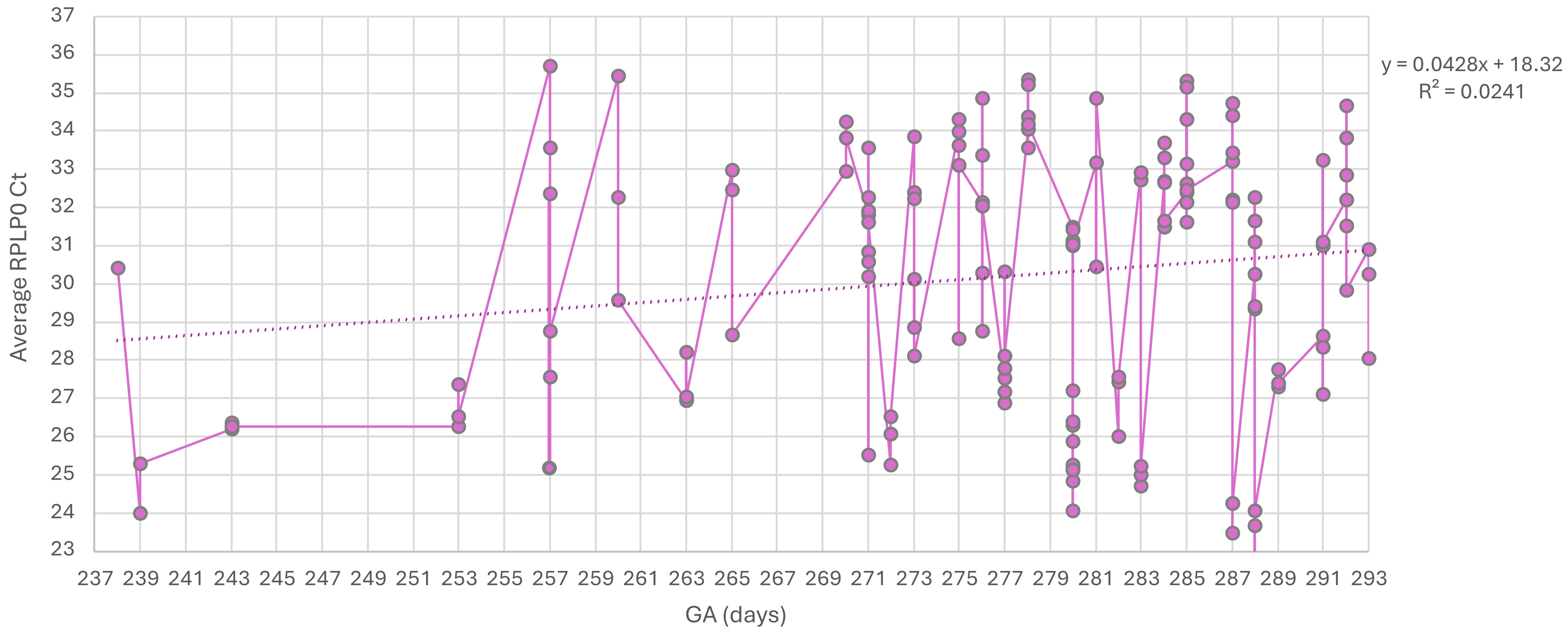

### Additional File 8

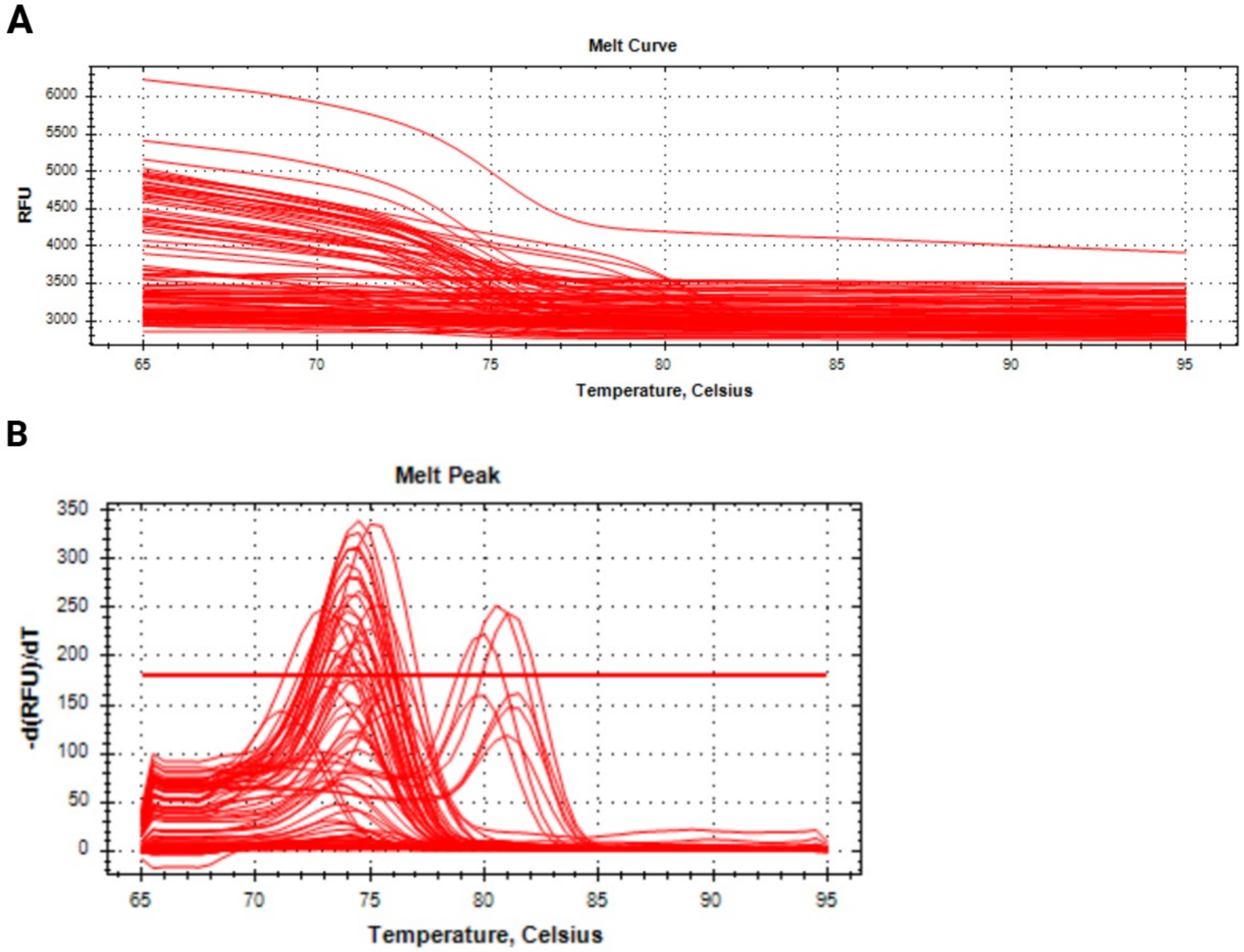

### Additional File 9

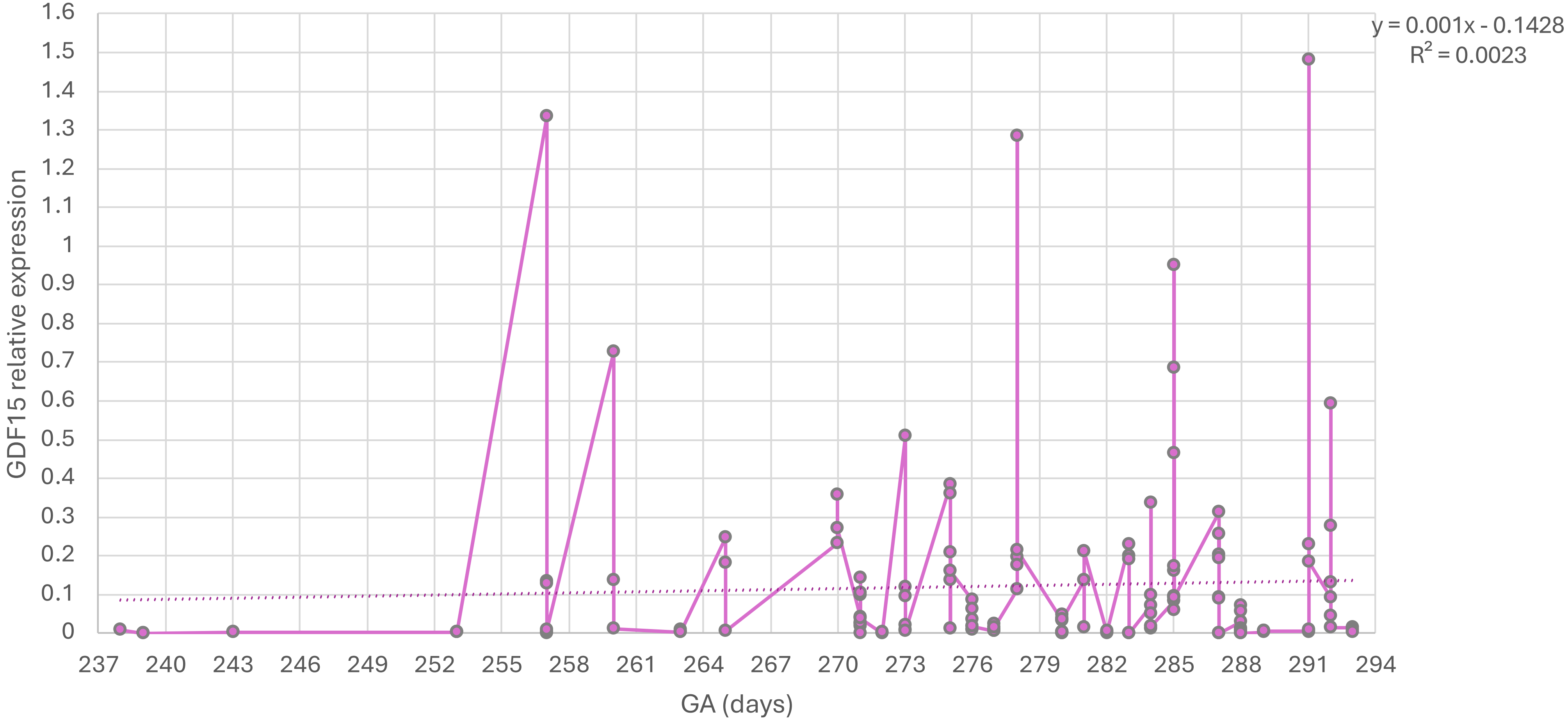

### Additional File 10

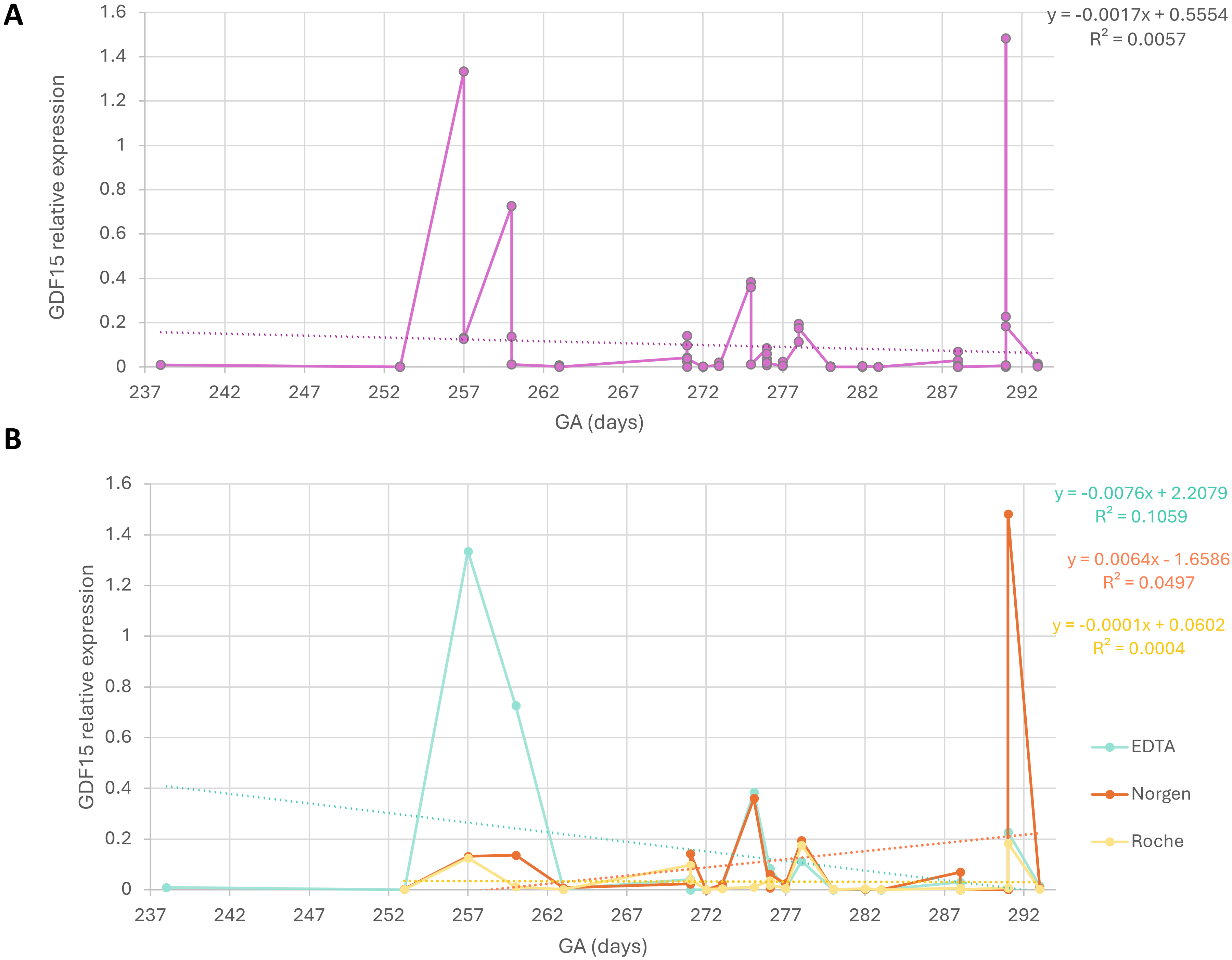
